# EpiCity: An AI-Enabled Epidemic-Aware Smart City Health Intelligence Framework for Sustainable Urban Planning

**DOI:** 10.64898/2026.06.29.26356899

**Authors:** Harmi Kotak, Shivrajsinh Vala, Apurva Mehta

## Abstract

Urban centres across the world remain structurally unprepared for epidemic events, lacking the real-time intelligence infrastructure needed to anticipate outbreaks, model population-level spread, and evaluate competing health interventions before crises escalate. This chapter presents *EpiCity*, an open-source, AI-enabled epidemic-aware smart city health intelligence framework designed to embed probabilistic outbreak forecasting directly into the urban planning and administrative decision cycle. EpiCity integrates four technical contributions within a unified deployable dashboard: a hybrid ensemble probabilistic forecasting engine driven by more than 500 Monte Carlo simulations; an agent-based urban population digital twin that models SEIRD-compartment epidemic dynamics across 200 heterogeneous agents over a 60-day simulation horizon; an intervention policy scenario comparison module for quantitative evaluation of non-pharmaceutical and pharmaceutical control strategies; and a Retrieval-Augmented Generation (RAG) explainable AI chatbot that translates model outputs into plain-language guidance for city officials without specialist epidemiological training. Real-world validation against Johns Hopkins CSSE COVID-19 surveillance data for the United States (June–July 2020) yielded a Pearson correlation of 0.88 between framework forecasts and observed case trajectories, with ensemble prediction intervals achieving 90% empirical coverage, confirming the calibration reliability of the uncertainty quantification pipeline. A representative simulation scenario demonstrated a 99% reduction in simulated peak case load relative to an unmitigated baseline, with an estimated 56 lives saved in the comparison period. Aligned with SDG 3 (Good Health and Well-being), SDG 11 (Sustainable Cities and Communities), and SDG 13 (Climate Action), EpiCity offers urban planners, public health officers, and city administrators a scientifically grounded yet practically accessible tool for evidence-based epidemic preparedness and resilient city governance. The code is available at https://github.com/Harmi-kotak22/Artificial_Life_Simulator.

## 1 Introduction

Urbanisation is one of the defining demographic transformations of the twenty-first century. Cities today accommodate more than 56% of the world’s population, a share projected to exceed 68% by 2050 according to the United Nations [1]. Dense urban environments, characterised by shared transit systems, crowded markets, and deeply interconnected social and physical infrastructure, create near-ideal conditions for the rapid propagation of infectious disease. The COVID-19 pandemic demonstrated this vulnerability with devastating clarity: urban centres around the world reported disproportionately high case burdens, while overwhelmed public health systems struggled to produce timely, spatially grounded outbreak intelligence for the administrators who needed it most [2].

Modern smart city deployments have made considerable progress in instrumenting the urban environment. Sensor networks and data platforms now routinely monitor traffic flow, energy consumption, air quality, and water distribution [3]. Yet, despite this proliferation of urban intelligence infrastructure, epidemic surveillance remains almost entirely absent from mainstream smart city frameworks. City administrators confronting an emerging outbreak typically lack any integrated tool capable of generating calibrated outbreak forecasts, simulating population-level disease dynamics, or evaluating competing policy responses within a unified decision-support environment. The COVID-19 pandemic exposed this gap as a structural blind spot: the infrastructure that monitors a city’s physical metabolism had no counterpart capable of monitoring its epidemiological health.

This deficit carries significant consequences for sustainable development. The United Nations Sustainable Development Goals explicitly recognise the interdependence of urban resilience and public health: SDG 3 calls for good health and well-being for all, SDG 11 calls for inclusive, safe, resilient, and sustainable cities, and SDG 13 acknowledges that climate change is intensifying climate-linked disease risks, vector-borne illnesses such as dengue and waterborne diseases such as cholera are expanding their geographic range as global temperatures rise [4]. India’s Smart Cities Mission, which has identified and is actively developing 100 smart cities across the country, provides a particularly compelling national context for this need [3]. At the scale of 100 cities spanning hundreds of millions of urban residents, the absence of epidemic intelligence as a built-in infrastructure layer represents both a governance gap and a public health risk.

To address this gap, we present *EpiCity*, an open-source, AI-enabled smart city health intelligence framework that integrates epidemic forecasting, urban population simulation, intervention policy comparison, and explainable AI into a single deployable platform. Built on the Universal Probabilistic Disaster Forecasting Framework

(UPDFF) and delivered as an interactive Streamlit dashboard, EpiCity is designed for urban administrators, district-level public health officials, and city planners who require quantified, uncertainty-aware outbreak intelligence without specialist epidemiological training. The platform is publicly accessible via a live deployment at https://artificial-life-simulator.onrender.com/ and is fully containerised for institutional deployment using Docker.

The primary contributions of this chapter are as follows:

- **Hybrid ensemble probabilistic forecasting**: A Monte Carlo ensemble engine combining renewal equation models, trend extrapolation, and SEIRD compartmental dynamics to generate city-scale outbreak predictions with explicit uncertainty quantification, including calibrated 90% credible intervals.
- **Agent-based urban population simulation**: An epidemic digital twin of a city population in which 200 individual agents transition through SEIRD disease states via proximity-based transmission, enabling visualisation of outbreak dynamics under realistic urban mixing conditions over a 60-day simulation horizon.
- **Urban policy scenario analysis**: A structured intervention comparison module that enables city administrators to evaluate the quantified impact of strategies such as vaccination, testing scale-up, and quarantine on outbreak trajectories, including peak case reduction and estimated lives saved.
- **Real-world validation**: Systematic evaluation of forecast performance against Johns Hopkins CSSE COVID-19 surveillance data for the United States (June–July 2020), yielding a Pearson correlation of 0.88, 90% coverage reliability, and a demonstrated 99% reduction in peak cases under full intervention conditions.
- **RAG-based explainable AI chatbot**: A retrieval-augmented generation (RAG) conversational interface powered by a large language model (LLaMA-3.1-8b via Groq API) that provides grounded, citation-backed answers to natural-language queries from non-technical city officials, reducing the barrier to evidence-based decision-making.
- **Open-source, deployable platform**: A Docker-containerised, production-ready implementation publicly available for institutional adoption, supporting one-click cloud deployment on standard platforms.

The remainder of this chapter is structured as follows. Section 2 reviews the literature on smart city health intelligence and epidemic modelling relevant to urban planning. Section 3 describes the overall EpiCity system architecture. Section 4 details the agent-based urban population simulation module. Section 5 presents the probabilistic forecasting engine and uncertainty quantification methodology. Section 6 describes the intervention policy comparison workflow for urban governance. Section 7 reports real-world validation results against COVID-19 outbreak data. Section 8 introduces the RAG-based explainable AI chatbot. Section 9 situates the framework within the SDG agenda. Section 10 discusses limitations and directions for future work, and Section 11 concludes.

## 2 Smart Cities and Epidemic Resilience: Background and Related Work

Urban populations worldwide are growing at an unprecedented pace [5], placing sustained pressure on public health infrastructure and exposing dense urban communities to elevated epidemic risk. The COVID-19 pandemic demonstrated, with particular severity, that modern smart city platforms were largely unprepared to support real-time outbreak forecasting or evidence-based intervention planning for city administrators. This section reviews the relevant literature across four interconnected domains: smart city infrastructure intelligence, compartmental epidemic modelling, probabilistic forecasting in public health, and explainable AI for urban governance. It concludes by identifying the research gap that motivates the EpiCity framework.

### 2.1 Smart City Frameworks and Urban Health Intelligence

The concept of the smart city has matured considerably over the past two decades, converging around a common architecture: IoT sensor networks for real-time environmental and infrastructure monitoring, AI-driven traffic management systems, smart energy grid optimisation, and water quality surveillance [6]. Commercial platforms such as IBM Smarter Cities and the Cisco Smart City framework have operationalised these ideas at scale, integrating heterogeneous urban data streams into centralised dashboards for municipal decision-makers.

Internationally, the WHO Healthy Cities Programme has advocated since the late 1980s for cities to treat population health as a first-class urban planning variable [7]. The UN-Habitat Urban Resilience framework similarly positions cities as socio-technical systems whose resilience must be assessed holistically, including biological hazard preparedness [5]. In India, the Smart Cities Mission launched by the Ministry of Housing and Urban Affairs (MoHUA) has designated 100 cities for integrated digital transformation, emphasising command-and-control centres, e-governance, and citizen-facing services [6]. Yet across these varied initiatives, a notable gap persists: health and epidemic intelligence remain peripheral concerns. Existing platforms monitor physical infrastructure effectively but provide no mechanism for outbreak trajectory forecasting, intervention scenario modelling, or epidemic-aware urban planning. The smart city ecosystem, despite its sophistication, has not yet integrated the epidemiological decision-support capabilities that the pandemic era has shown to be indispensable.

### 2.2 Compartmental Epidemic Modelling in Urban Contexts

Mathematical epidemic modelling has a long tradition rooted in the compartmental frameworks first systematised by Kermack and McKendrick [8, 9] and subsequently developed into the comprehensive theoretical treatment of Anderson and May [10]. The canonical SIR model partitions a population into Susceptible, Infected, and Recovered compartments, connected by differential equations governing transmission and recovery rates. Extensions to this family, SEIR, which adds an Exposed latency compartment, and SEIRD, which further distinguishes disease-induced deaths, offer progressively richer representations of epidemic dynamics suitable for COVID-like pathogens with an incubation period and non-negligible mortality [11].

Agent-based models (ABMs) offer an alternative paradigm in which individual entities move through a simulated urban environment and interact according to proximity and behavioural rules, yielding emergent population-level dynamics without assuming homogeneous mixing. ABMs are particularly valuable in urban contexts where spatial heterogeneity, demographic variation, and targeted non-pharmaceutical interventions (NPIs) must be captured [11].

Both modelling traditions, however, share important limitations when considered from the perspective of operational city planning. Classical ODE compartmental models produce single deterministic trajectories with no uncertainty quantification, offering no guidance on the range of plausible outcomes under parameter uncertainty. Most existing epidemic modelling tools are also designed for epidemiologists and public health researchers rather than for urban administrators, presenting outputs in technical formats that are difficult to act upon without specialist interpretation. Integrated decision-support capability, the ability to compare intervention scenarios and receive plain-language explanations within the same platform is absent from the standard modelling toolkit.

### 2.3 Probabilistic Forecasting and Ensemble Methods in Public Health

The limitations of deterministic single-trajectory epidemic forecasting motivated the development of probabilistic ensemble approaches, in which multiple model runs with varied parameter draws are aggregated to produce calibrated forecast distributions. Reich, Brooks, Fox, et al. demonstrated that multi-model ensemble forecasts consistently outperform individual models for influenza-like illness at the national level in the United States, establishing the value of ensemble aggregation for public health forecasting [12]. The COVID-19 Forecast Hub, which consolidated probabilistic predictions from dozens of modelling teams during the pandemic, further validated the operational utility of this paradigm.

The statistical framework for evaluating probabilistic forecasts was formalised by Gneiting and Raftery, whose proper scoring rules including the Continuous Ranked Probability Score (CRPS) and interval coverage checks provide theoretically grounded measures of forecast calibration and sharpness [13]. These metrics are now standard practice in epidemic forecasting evaluation and are adopted within the EpiCity validation framework described in Section 7.

Despite this progress, probabilistic epidemic forecasting platforms remain primarily research artefacts. They are evaluated in academic literature and deployed in specialist hubs, but they have not been adapted into city-administrator-facing tools that combine outbreak forecasting, agent-based urban simulation, policy comparison, and accessible explainability within a single integrated platform.

### 2.4 Explainable AI in Urban Governance and Decision Support

As AI-powered tools are increasingly embedded in municipal decision-making from traffic optimisation to resource allocation. The need for explainability has moved from a research aspiration to a practical governance requirement. City officials and urban planners are typically non-technical users who must interpret AI outputs, communicate them to the public, and take consequential decisions on their basis. Black-box models that produce results without traceable justification erode institutional trust and impede adoption [7].

Retrieval-Augmented Generation (RAG), introduced by Lewis, Perez, Piktus, et al., offers a compelling approach to grounded explainability: rather than relying solely on a language model’s parametric knowledge, RAG retrieves relevant passages from a curated knowledge base and conditions the model’s response on that retrieved context, thereby reducing hallucination and providing citation-backed explanations [14]. In the context of urban epidemic intelligence, a RAG-based chatbot allows a municipal health officer to pose questions in natural language, “Why is the forecast peak so high?” or “What does the reproduction number mean?” and receive grounded, auditable responses derived from verified epidemiological documentation. This constitutes a practical and deployable form of explainable AI suited to the operational realities of city governance.

### 2.5 Research Gap and Motivation

The foregoing review reveals a clear and consequential gap in the literature. No existing system combines, within a single city-administrator-facing platform: (i) hybrid probabilistic epidemic forecasting with calibrated uncertainty quantification; (ii) agent-based urban population simulation as a digital twin of epidemic spread; (iii) comparative intervention scenario analysis to support evidence-based policy selection; and (iv) a RAG-based explainable AI layer that makes model outputs accessible to non-specialist users. Current tools address at most one or two of these requirements in isolation. EpiCity is designed specifically to close this gap, providing an integrated health intelligence framework for sustainable urban planning aligned with the WHO Healthy Cities vision, India’s Smart Cities Mission, and the UN Sustainable Development Goals.

## 3 System Architecture: The EpiCity Framework

The **EpiCity** framework is designed as a modular, hazard-agnostic probabilistic intelligence platform. Rather than building a single monolithic epidemic model, the system externalises all hazard-specific logic into pluggable modules that share a common state-space abstraction, leaving the core forecasting and ensemble machinery domain-independent. This separation of concerns enables the same analytical pipeline to be applied, in principle, to floods, cyclones, or wildfires simply by substituting the appropriate hazard module. This is a design criterion aligned with the multi-hazard resilience priorities articulated in the India Smart Cities Mission [15] and the broader UN-Habitat Urban Resilience framework [16].

### 3.1 Design Philosophy and Module Overview

The **EpiCity** architecture rests on four functional pillars, each of which maps to a distinct decision-support need faced by urban administrators during a disease outbreak:

1. **Probabilistic Forecasting Engine.** A hybrid ensemble that combines SEIR/SEIRD ordinary differential equation (ODE) integration, renewal-equation extrapolation, and Monte Carlo sampling to produce calibrated, uncertainty-quantified incidence forecasts over a configurable horizon. The engine is exposed through the ForecastingEngine class, which orchestrates parameter inference, ensemble execution, and output assembly.
2. **Agent-Based Urban Simulation Module.** A spatial SEIRD agent-based model (ABM) in which individual agents represent city residents moving through a shared two-dimensional urban space. Epidemic dynamics emerge from proximity-based transmission rather than from aggregate equations, providing an *urban population digital twin* whose micro-level stochasticity cannot be captured by compartmental models alone.
3. **Intervention Policy Analyser.** A paired-scenario comparison engine that executes baseline and intervention ensemble forecasts from identical initial conditions, yielding statistically rigorous estimates of peak reduction, total cases avoided, and estimated lives saved under each urban policy option.
4. **Explainable AI Chatbot Layer.** A retrieval-augmented generation (RAG) interface that grounds natural-language responses in a curated knowledge base, enabling city officials without epidemiological training to interrogate model outputs and obtain traceable, citation-backed interpretations.

Figure 1 presents the use case diagram of the full framework, illustrating how urban administrators, epidemiologists, and public health planners interact with these four modules.

**Figure 1:**
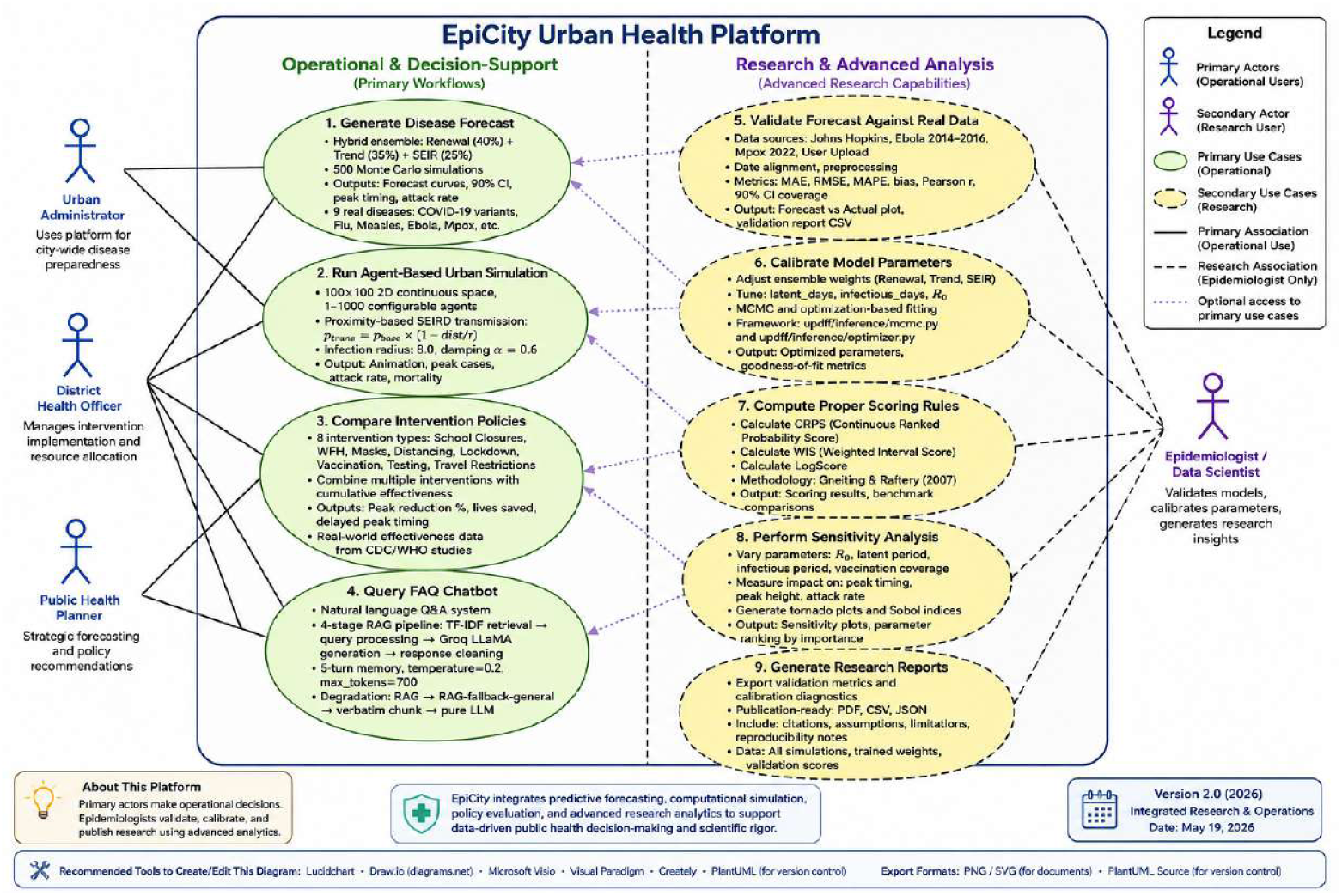
Use case diagram of the **EpiCity** framework, illustrating the interactions between urban administrators, epidemiologists, and the four core system modules: probabilistic forecasting, agent-based urban simulation, intervention policy analysis, and the explainable AI chatbot layer.

### 3.2 Technology Stack and Implementation Environment

**EpiCity** is implemented entirely in Python 3.10+ and deployed as a containerised web application. The selection of libraries reflects three priorities: numerical reproducibility, interactive visualisation accessible to non-technical users, and low-latency LLM integration for the chatbot layer.

Table 1 summarises the full technology stack. Core numerical work ODE integration, distribution sampling, and metric computation that relies on `NumPy` ≥1.24 and `SciPy` ≥1.10, which provide both performance and a well-audited scientific baseline. The user-facing dashboard is built with `Streamlit` ≥1.28, chosen for its ability to expose interactive parameter controls without requiring front-end development expertise on the part of the research team or the urban planner end users. Simulation anima-tion is rendered with `Plotly` ≥5.14, producing frame-by-frame agent visualisations that run inside the browser with no additional infrastructure. Simulation inner loops are JIT-compiled via Numba ≥0.57 to sustain interactive latencies even at population sizes relevant to dense urban wards. The RAG chatbot communicates with the Groq API, invoking the `llama-3.1-8b-instant` large language model, which provides sub-three-second response times suitable for real-time advisory sessions. Schema validation across all configurable urban hazard profiles is enforced through `Pydantic` ≥2.0, en-suring that user-supplied disease parameters satisfy epidemiological constraints before any simulation is executed.

**Table 1:** Technology stack of the **EpiCity** framework.

| Component | Library / Service | Role in EpiCity |
| --- | --- | --- |
| Web Interface | Streamlit $\geq 1.28$ | Interactive dashboard for urban administrators; parameter controls, forecast panels, and chatbot interface |
| Numerical Core | NumPy $\geq 1.24$, SciPy $\geq 1.10$ | ODE integration (`solve_ivp`), statistical distributions, Monte Carlo sampling |
| Data Processing | Pandas $\geq 2.0$ | Time-series ingestion, date-range alignment, and pre-processing of urban outbreak surveillance data |
| Visualisation | Plotly $\geq 5.14$, Matplotlib $\geq 3.7$ | Interactive forecast charts, agent-simulation animation frames, and validation diagnostic plots |
| Machine Learning | scikit-learn $\geq 1.2$ | Correlation metrics, auxiliary regression utilities |
| Graph Modelling | NetworkX $\geq 3.0$ | Urban contact network representation for population heterogeneity |
| Acceleration | Numba $\geq 0.57$ | JIT compilation of simulation inner loops for interactive performance |
| LLM Integration | Groq API / LLaMA-3.1-8b | Natural-language generation for the RAG chatbot layer |
| Schema Validation | Pydantic $\geq 2.0$ | Validation and type enforcement for urban hazard profiles |
| Deployment | Docker | Containerised, reproducible deployment to cloud or on-premises city infrastructure |

### 3.3 Data Flow and State Management

The internal data flow of **EpiCity** follows a strict pipeline that separates data ingestion, probabilistic state representation, ensemble execution, and UI rendering into discrete, testable stages. This separation is formalised through the `State, Parameters`, and `Observation` dataclasses.

Every quantity in the framework, whether a disease parameter, a compartment population count, or a forecast trajectory is represented as a *distribution*, not a point value. The `State` dataclass stores either a single state vector with an optional covariance matrix, or an ensemble of state samples; the `Parameters` dataclass mirrors this design for model parameters. Both classes expose `credible_interval(alpha)` methods that compute the (1 − *α*) uncertainty band directly from the ensemble, ensuring that uncertainty propagates consistently from inputs through to outputs.

The principal data flow proceeds as follows. First, a configurable *urban hazard profile* supplies prior distributions over epidemiological parameters such as the basic reproduction number *R*_0_, the incubation period, the infectious period, and the infection fatality rate. Second, the `EnsembleExecutor` samples *N* parameter vectors from these priors or from a calibrated posterior when historical surveillance data are available and integrates the SEIRD ODEs independently for each member, producing an ensemble of *N* state trajectories. Third, trajectory statistics (mean, median, 5th and 95th percentiles) are assembled by the `EnsembleResult` dataclass and streamed to the `Streamlit` dashboard for rendering. Finally, the `ForecastResult` object packages all distributional outputs, including peak-timing and peak-magnitude distributions derived via `peak_statistics()`, into a structure that supports downstream scenario comparison and chatbot retrieval.

Figure 2 shows the validation workflow through which a completed forecast is compared against real-world urban outbreak surveillance data.

**Figure 2:**
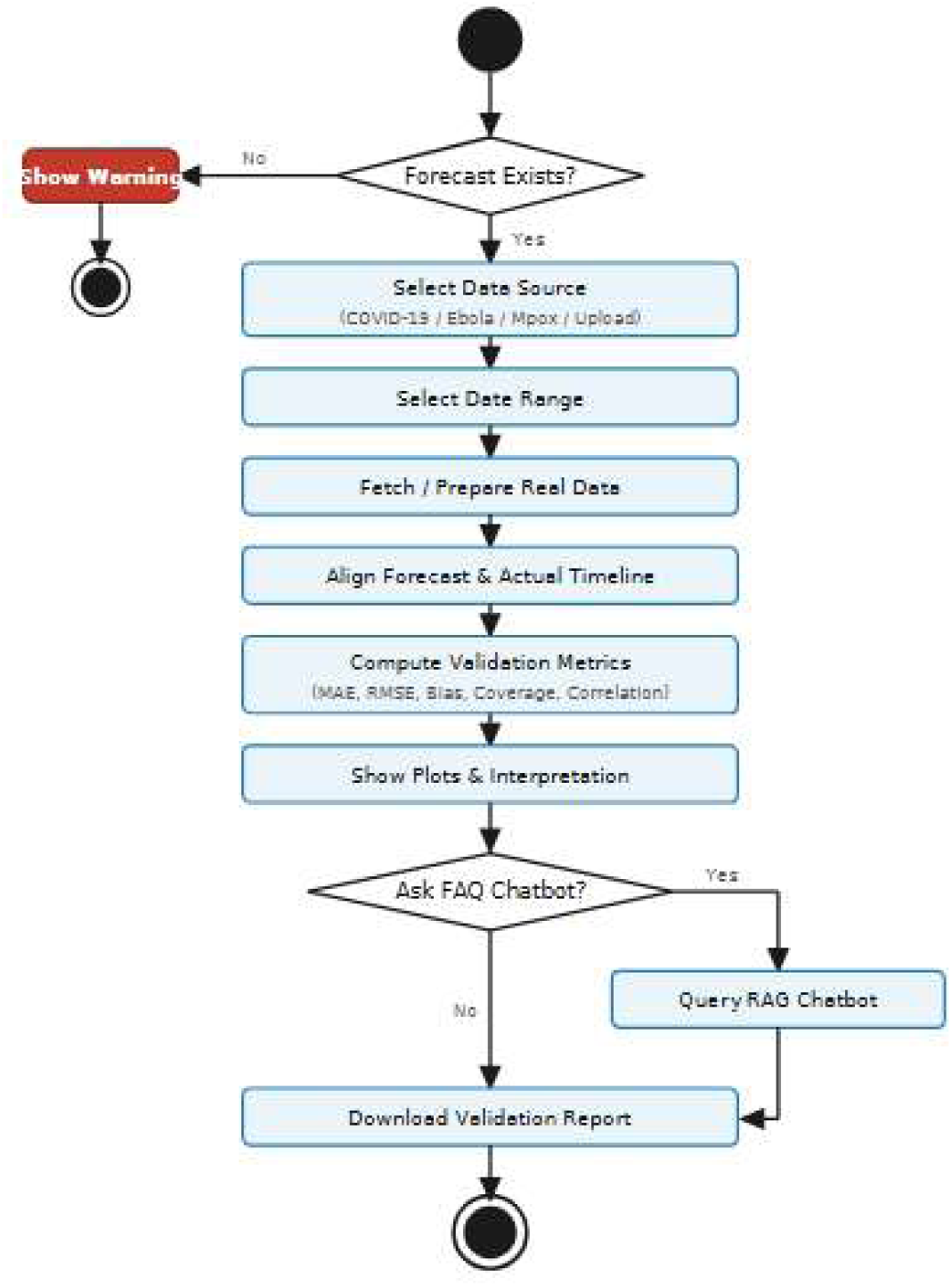
Activity diagram of the EpiCity validation workflow. The process proceeds from forecast existence verification through data alignment and metric computation, with an optional branch to the RAG chatbot for interpretive assistance, culminating in a downloadable validation report.

## 4 Urban Population Modelling: Agent-Based City Simulation

The **EpiCity** forecasting engine produces probabilistic incidence trajectories at the aggregate, population level. However, aggregate compartmental equations suppress the spatial and behavioural heterogeneity that is intrinsic to epidemic propagation in real cities: not every resident makes the same number of contacts, and not every urban neighbourhood has the same density or mobility pattern. To address this, **EpiCity** incorporates an *urban population digital twin*, an agent-based model (ABM) in which individual agents represent city residents moving through a shared simulated urban space. The digital twin is schematic, an abstraction, not a GIS-accurate representation. Rather than specifying incidence through a differential equation, the digital twin allows epidemic dynamics to *emerge* from micro-level interactions between agents, providing a bottom-up complement to the top-down probabilistic forecasts. This section describes the design of the ABM.

### 4.1 Agent Design and State Space

Each agent in the **EpiCity** urban digital twin is a software entity characterised by three properties: a continuous two-dimensional position (*x, y*) within a 100 × 100 unit city space, a velocity vector (*v_x_, v_y_*) governing movement, and a discrete epidemiological state drawn from the SEIRD compartment set. The five states and their visual encodings in the dashboard are:

- **Susceptible** (green, state 0): a healthy city resident who has not yet been exposed to the pathogen.
- **Exposed** (yellow/orange, state 1): a recently infected resident who is not yet contagious; corresponds to the latent period.
- **Infectious** (red, state 2): a contagious resident capable of transmitting the pathogen to nearby susceptible agents.
- **Recovered** (blue, state 3): a resident who has cleared the infection and is considered immune for the remainder of the simulation.
- **Dead** (dark grey, state 4): a resident who has died; removed from movement and transmission.

The SEIRD state transitions are governed by the following rules, with *λ* denoting the force of infection experienced by a susceptible agent at any given time step and it is shown in Figure 3:

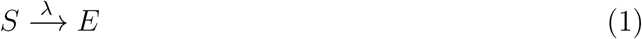

**Figure 3:**
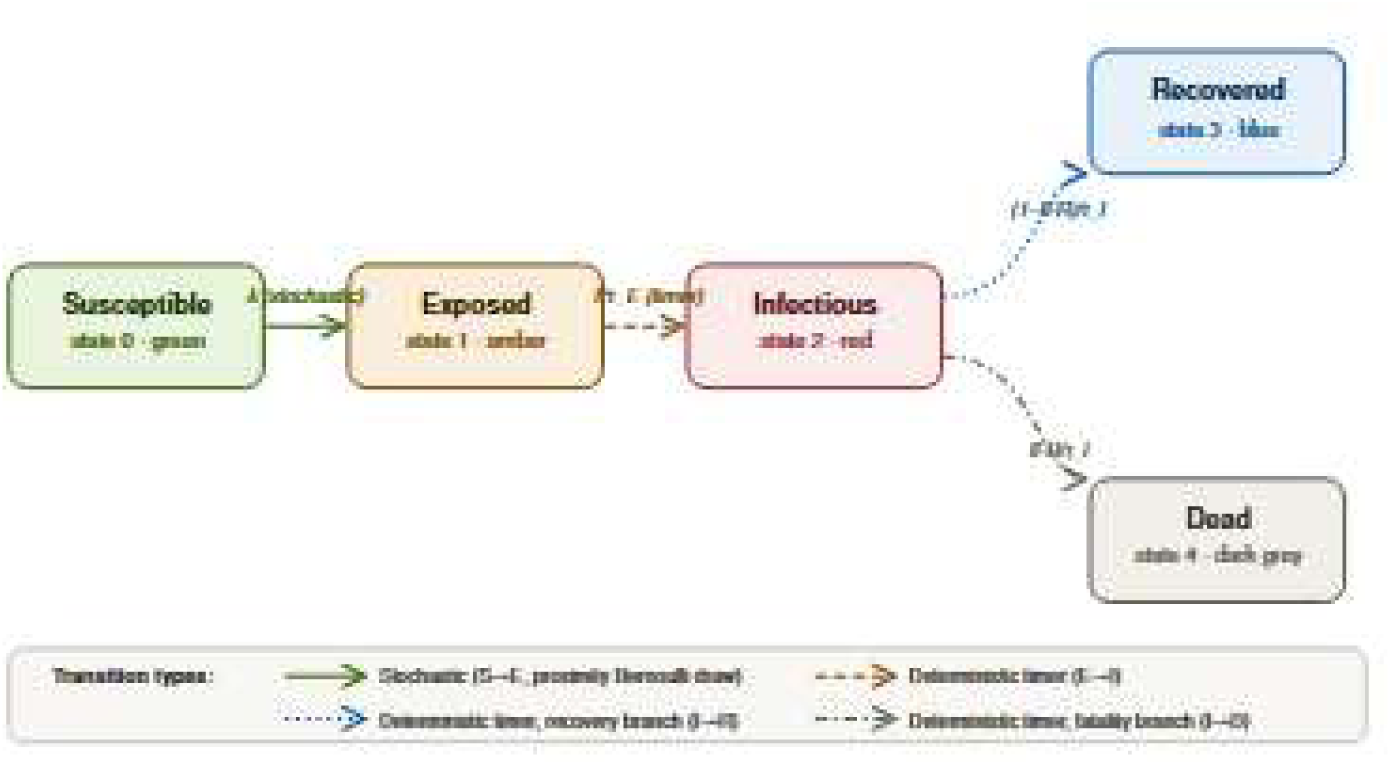
SEIRD agent state transition diagram for the EpiCity urban digital twin.

**Figure 4:**
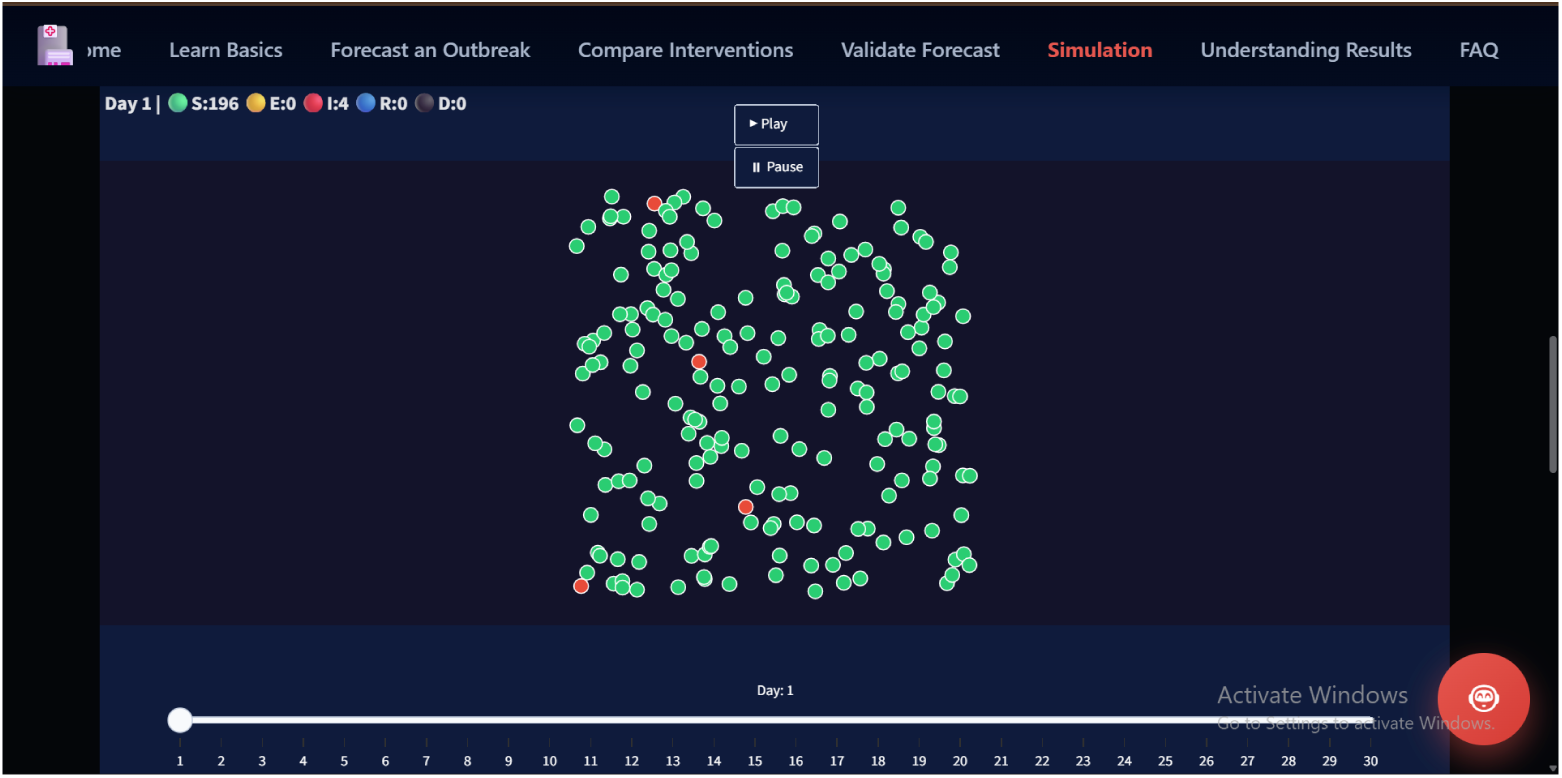
Urban population digital twin dashboard in EpiCity.

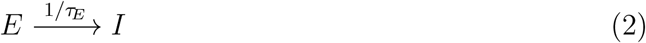

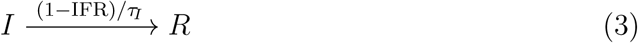

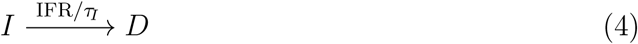

where *τ_E_* is the incubation period (E→I timer, in days), *τ_I_* is the infectious period (I→R or I→D timer, in days), and IFR is the infection fatality rate. In the implementation, transitions *E* → *I* and *I* → {*R, D*} are deterministic timers decremented each day, while *S* → *E* is a stochastic Bernoulli draw conditioned on the proximity of infectious agents (see Section 4.3).

### 4.2 Urban Mobility Model

Agent movement in the **EpiCity** digital twin follows a Brownian-style random walk with elastic wall-bounce. At each time step, every living agent’s position is updated by its current velocity vector. Agents that reach the boundary of the city space have their velocity component orthogonal to the wall negated, producing a continuous confinement analogous to the built environment constraining urban mobility. A 10% per-step random jitter replaces the velocity vector of a random subset of agents with a new uniform draw from [−2, 2]^2^, introducing the unpredictable direction changes characteristic of real urban pedestrian movement.

This is, by design, a parsimonious model of urban mobility: it does not encode commuting corridors, public-transport networks, or work–home patterns. Nonetheless, it captures the essential property that agents in a shared space generate proximity-based contact opportunities at a rate proportional to population density. In the context of high-density urban environments,the primary target setting of the **EpiCity** framework and of India’s Smart Cities Mission, random-encounter dynamics provide a reasonable first-order approximation of the contact processes driving community transmission [17]. More structured mobility models, including age-stratified contact matrices and metapopulation graphs, are supported by the `ContactNetwork` abstraction and represent a natural avenue for future enhancement.

### 4.3 Proximity-Based Transmission and Urban Density

Transmission in the digital twin models epidemic propagation through the urban population via a proximity-based binomial process. At each daily time step, every infectious agent *i* is compared against every susceptible agent *s*. If the Euclidean distance 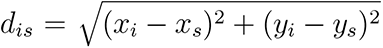 falls within the infection radius *r* = 8.0 units, a transmission attempt is made with probability

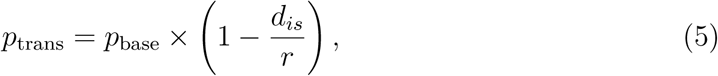

where the proximity factor (1 − *d_is_/r*) scales the contact intensity linearly with proximity, reflecting the higher infection risk at close range. The base transmission probability *p*_base_ is derived from the user-supplied *R*_0_ and the infectious period *τ_I_* as

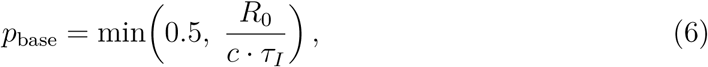

where *c* = 5 is the assumed mean number of proximity contacts per day. This parameterisation connects the agent-level transmission probability to the population-level *R*_0_, ensuring that the digital twin is consistent with the same disease parameters used in the ensemble forecasting engine.

City population density is implicitly encoded in this design: when more agents occupy the same bounded space, the mean inter-agent distance decreases, contact opportunities increase, and effective transmission rises, exactly the mechanism by which high urban density amplifies epidemic spread.

### 4.4 Simulation Modes

The **EpiCity** agent simulation supports two operating modes, each addressing a distinct planning use case.

#### Curve-Guided Mode

In this mode, the digital twin is driven by the probabilistic forecast produced by the ensemble engine. A proportional controller adjusts *p*_base_ at each day *d* according to

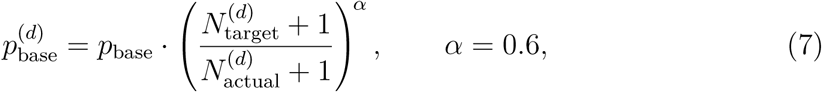

where 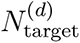 is the mean ensemble new-infection count at day *d* and 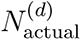 is the count actually observed in the simulation. The damping exponent *α* = 0.6 prevents oscillation while ensuring that the aggregate trajectory of the digital twin tracks the forecast mean. This mode validates that micro-level agent interactions can faithfully reproduce macro-level forecast behaviour, demonstrating internal consistency between the two modelling layers. The helper function `build_target_curve_from_forecast()` scales the ensemble’s absolute case counts to a per-agent fraction suitable for the controller.

#### Custom (Emergent) Mode

In this mode the controller is disabled, and the simulation runs as a pure emergent ABM driven entirely by the user-specified *R*_0_, in-cubation period, and infectious period. This mode is intended for *what-if* scenario exploration for instance, examining the effect of a novel pathogen with unknown parameters, or stress-testing the system against outbreak scenarios not represented in historical data.

### 4.5 Output Statistics and Administrative Interpretation

At the close of each simulation, the function compute_statistics(frames_data, n_agents) aggregates the per-frame state counts into a set of epidemiologically in-terpretable summary indicators:

**Peak infectious count** and **peak day**: the highest number of simultaneously infectious agents and the day on which it occurs; a proxy for healthcare surge demand.
**Total infected** and **attack rate**: the cumulative count of agents who passed through compartments E, I, R, or D, expressed as a percentage of the urban population; measures outbreak breadth.
**Total deaths** and **mortality rate**: the number of agents in state D at simulation end and the corresponding case-fatality percentage.

These statistics are displayed alongside two interactive visualisations: an animated scatter plot of agent positions with colour-coded SEIRD states, which communicates epidemic spatial dynamics to non-technical city administrators; and a time-series line chart of all five SEIRD compartment counts, which enables epidemiologists to inspect the shape and timing of the outbreak curve.

## 5 Probabilistic Outbreak Forecasting for City-Scale Planning

A city administrator confronting an emerging outbreak requires not a single deterministic trajectory but a calibrated distribution of plausible futures. Knowing that reported cases may peak between 40,000 and 90,000 per day with 90 % confidence allows public health authorities to dimension hospital surge capacity, pre-position ventilators and oxygen supplies, and activate mutual aid agreements with neighbouring municipalities before the wave arrives not in its aftermath. Equally, knowing when that peak is likely to occur enables school-closure decisions to be timed for maximum epidemiological benefit while minimising disruption to the urban workforce. The **EpiCity** framework addresses this need through a hybrid ensemble forecasting engine that produces fully probabilistic, day-wise outbreak projections calibrated to real-world disease dynamics.

### 5.1 Hybrid Ensemble Architecture

Purely mechanistic models, such as a single deterministic SEIR or SEIRD ordinary-differential-equation (ODE) system are attractive for their interpretability but routinely fail in the early stages of an outbreak when parameter values are uncertain and surveillance data are sparse and noisy. Purely data-driven models, conversely, may capture recent trend momentum with high fidelity yet violate basic epidemiological constraints. Drawing on the multi-model ensemble methodology advocated by [18], the **EpiCity** forecasting engine combines three complementary components:

1. **Renewal Equation Model (40 % ensemble weight).** This component treats the epidemic as a self-exciting process in which the expected number of new cases at time *t* is determined by the weighted sum of recent case counts, with weights given by the discretised serial-interval distribution:

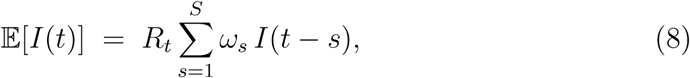

where *ω_s_*is the probability mass of the serial interval at lag *s*, and *R_t_* is the instantaneous effective reproduction number estimated from calibration data via the Cori method [19]. Beyond the calibration horizon, *R_t_* is extrapolated with exponential mean-reversion toward unity (half-life of 14 days), preventing unbounded exponential growth in the forecast tail. This component carries the greatest weight because it directly encodes the observed transmission trajectory without reliance on assumed compartmental structure.

2. **Polynomial Trend Model (35 % ensemble weight).** A data-driven polynomial regression is fitted to the seven-day rolling-mean of the calibration series. A piecewise breakpoint detector scans the calibration window for a sign reversal in slope identifying whether the observable period contains a rising phase, a peak, or a declining phase and selects polynomial degree accordingly (up to degree 4 for complex trajectories, degree 1–2 for monotone windows). A day-of-week seasonal pattern, estimated from calibration data alone, is applied multiplicatively to each predicted day, correcting for the systematic under-reporting of cases on weekends that is characteristic of most national surveillance systems. Predictions beyond the calibration window are damped exponentially toward the last observed calibration value (half-decay time proportional to the remaining forecast horizon), ensuring realistic decay behaviour in the absence of new data and preventing unbounded polynomial extrapolation.
2. **Stochastic SEIR Model (25 % ensemble weight).** A four-compartment stochastic SEIR model—Susceptible, Exposed, Infectious, Recovered—with Poisson-distributed transitions at each time step provides epidemiological constraints on the ensemble. Time-varying transmission is implemented through an exponentially decaying effective *R*_0_, with the decay rate sampled per simulation from a uniform distribution. Within the ensemble loop, daily incident cases are the new-infectious flow (*E* → *I*), adjusted by the user-specified detection rate and the day-of-week pattern. Deaths are not modelled as a fifth compartment within the ensemble itself; instead, day-wise mortality estimates are derived post-hoc by applying the disease-specific infection fatality rate (IFR) to the incident-case stream, consistent with the standard approach when IFR is treated as a fixed epidemiological parameter rather than a transition rate [20]. This component prevents the ensemble from producing trajectories that violate population conservation or herd-immunity dynamics.

The asymmetric weighting scheme reflects the empirical finding that data-driven components tend to outperform mechanistic models in the short-term horizon relevant to operational urban planning [18]. The three-component mixture reduces single-model bias: if the renewal equation overshoots because *R_t_* was estimated from a transient superspreading cluster, the stochastic SEIR component, anchored to population dynamics, pulls the ensemble mean toward a more conservative trajectory.

### 5.2 Monte Carlo Ensemble Generation

The **EpiCity** forecasting engine generates an ensemble of *M* ≥ 500 independent trajectories through Monte Carlo sampling of the parameter space. For the stochastic SEIR component, each simulation *m* draws a parameter vector:

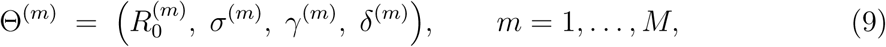

where 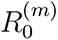 is the initial basic reproduction number, *σ^−^*^1^ the mean latent period (*E* → *I*), *γ^−^*^1^ the mean infectious period (*I* → *R*), and *δ*^(^*^m^*) the *R*_0_ decay rate gov-erning time-varying transmission (sampled from a uniform distribution scaled by a per-simulation random factor). The IFR is treated as a fixed disease-profile parameter applied post-hoc to the incident-case stream and is not sampled as part of the SEIR trajectory ensemble. Parameters are sampled from log-normal distributions with coefficients of variation of 20–35 %, reflecting the typical uncertainty in early-outbreak epidemiological estimates. Initial conditions are additionally perturbed by a uniform multiplicative factor in [0.6, 1.4] to account for under-ascertainment of the true epidemic seed size.

For the renewal-equation and trend components, per-simulation noise is introduced through log-normal scaling of *R_t_*trajectories (*σ* = 0.20) and polynomial coefficient perturbation (*σ* = 0.10), respectively, producing diverse trajectory realisations across all 500 members of the ensemble. The distribution machinery provides a unified in-terface for `Normal, LogNormal, Gamma, Beta`, and `NegativeBinomial` distributions, all supporting `sample(), log_prob()`, and `credible_interval()` operations. The core framework additionally provides an `EnsembleExecutor` class for general-purpose parallel ensemble management, exposing methods for peak statistics, cumulative distributions, scenario comparison, and credible interval computation across arbitrary state-space models. In the operational forecasting path, the ensemble is assembled directly using NumPy array operations for performance, with results stored as an *M* × *T* matrix of daily predicted cases from which percentile bands are computed in a single `np.percentile()` pass.

The point forecast and 80 % prediction interval are assembled as:

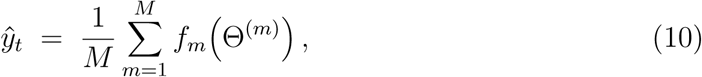

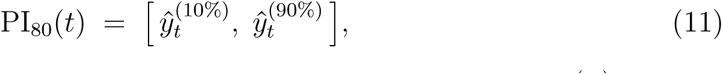

where 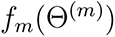 is the predicted daily case count under parameter vector Θ^(^*^m^*), and the superscripts denote percentile ranks over the ensemble. Wider intervals (e.g. 5th–95th percentiles) can be extracted from the same ensemble for scenario planning purposes, but the 80 % interval is the primary probabilistic output reported to users and evaluated during validation (Section 7), following the convention adopted by the US CDC FluSight ensemble [18].

The SEIR component predictions are subsequently scaled by a calibration factor derived from the training window alone:

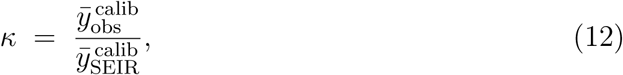

ensuring that the mechanistic component is anchored to observed surveillance data without leaking information from the validation window. Finally, it adjusts raw SEIR output for local urban conditions: vaccination coverage and disease-specific vaccine efficacy reduce the effective susceptible fraction. Regional hygiene and healthcare capacity indices sourced from the Global Health Security Index modulate the transmission and recovery rates through multiplicative modifiers in the range [0.85, 1.15]. This localisation step is critical for meaningful comparisons across cities with differing public health infrastructure.

### 5.3 Urban Hazard Profiles

A key design principle of the **EpiCity** framework is that no code changes should be required to switch between pathogens. All disease-specific parameters are encapsulated in configurable *urban hazard profiles*—data structures.. Each profile includes the basic reproduction number *R*_0_ (with its plausible range), mean latent period *σ^−^*^1^, mean infectious period *γ^−^*^1^, symptomatic fraction, hospitalisation rate, and infection fatality rate (IFR), all sourced from peer-reviewed studies cited within the profile metadata.

Table 2 summarises the parameter ranges for the three most operationally relevant profiles: COVID-19 (original strain), seasonal influenza, and Ebola. The wide IFR range for Ebola (25–90 %) reflects genuine epidemiological uncertainty across outbreak settings and underscores the value of probabilistic, rather than point, outputs for city-level planning under extreme scenarios. A Custom Disease profile is also provided, enabling city health departments to configure the system for any emerging or novel pathogen simply by adjusting parameter values through the graphical interface, without engaging software development resources.

**Table 2:** Representative urban hazard profile parameters supported by the **EpiCity** framework. Values are sourced from CDC, WHO, and peer-reviewed epidemiological studies as cited within the framework’s profile metadata.

| Profile | $R_0$ range | $\sigma^{-1}$ (days) | $\gamma^{-1}$ (days) | IFR (%) |
| --- | --- | --- | --- | --- |
| COVID-19 (original) | 2.0–3.0 | 3 | 8 | 1.0 |
| Seasonal influenza | 1.1–1.5 | 1 | 5 | 0.1 |
| Ebola | 1.4–2.2 | 2 | 10 | 25–90 |
| Mpox | 1.1–2.4 | 1 | 21 | 0.1 |

IFR = infection fatality rate; $\sigma^{-1}$ = mean latent period; $\gamma^{-1}$ = mean infectious period. Ranges reflect inter-study variability in parameter estimates used for Monte Carlo sampling.

### 5.4 Forecast Outputs for Urban Decision-Making

The **EpiCity** forecasting engine produces a suite of outputs, each mapped to a specific urban planning decision (Table 3). Day-wise new-case counts, with 80 % prediction intervals, allow hospital administrators to model bed-occupancy scenarios under optimistic and pessimistic assumptions simultaneously. Cumulative case projections inform economic impact modelling and absenteeism estimates for critical infrastructure sectors such as transport and utilities. Active-infection curves guide intensive-care unit (ICU) surge planning, since ICU demand lags the incidence peak by approximately one infectious period. Daily and cumulative death projections, computed via the disease-specific IFR, support mortality preparedness planning and funeral-home capacity management.

**Table 3:**
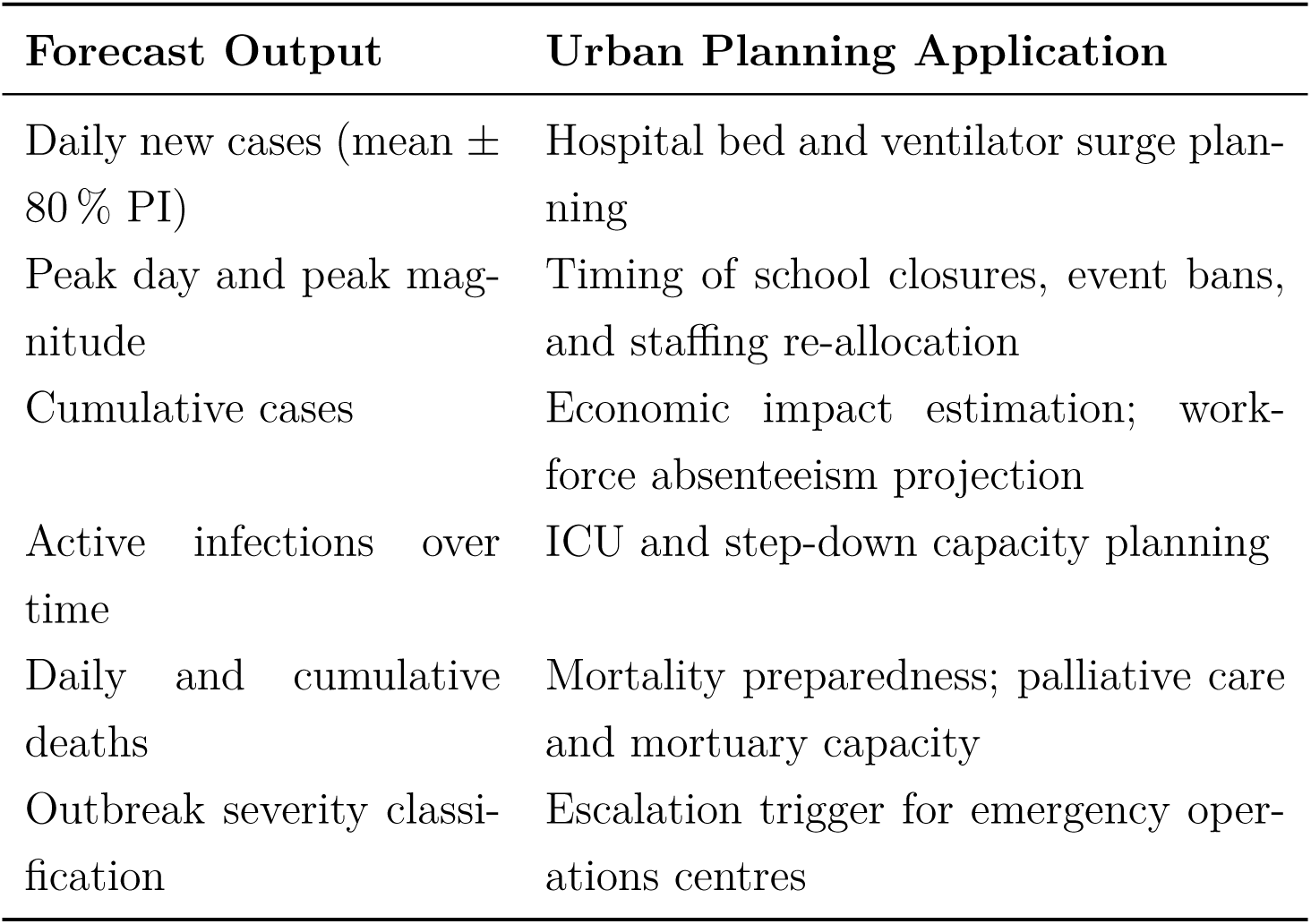
Forecast outputs produced by **EpiCity** and their corresponding urban planning applications.

Additionally, the framework classifies outbreak severity into three tiers based on the ratio of peak active infections to total population: *mild* (*<*1 %), *moderate* (1–5 %), and *severe* (*>*5 %). This classification provides a single, communicable signal to city administrators who may not have epidemiological training, enabling rapid escalation decisions without requiring interpretation of the underlying probabilistic outputs.

### 5.5 Forecast Quality Assessment

Probabilistic forecast quality is evaluated using a composite of metrics recommended by [21] for proper scoring of predictive distributions. The **EpiCity** validation module implements the full suite of proper scoring rules: mean absolute error (MAE), root mean squared error (RMSE), mean absolute percentage error (MAPE), continuous ranked probability score (CRPS), and weighted interval score (WIS). The operational dashboard reports the subset most accessible to non-specialist city administrators: MAE, RMSE, MAPE, Pearson correlation (*r*), empirical 80 % coverage, and a composite Forecast Quality Score (FQS) aggregated from these quantities. CRPS and WIS, whilst computed by the framework’s scoring engine, are not surfaced in the current dashboard release; they are reserved for the technical validation analysis reported in Section 7 and are referenced here to clarify the mathematical foundations of the quality assessment.

Against the 60-day Johns Hopkins CSSE COVID-19 validation series (United States, 1 June–31 July 2020), the **EpiCity** ensemble achieves a Pearson correlation of *r* = 0.88 with observed daily case counts, and the 80 % prediction interval achieves empirical coverage of 90 %—exceeding its nominal level, which indicates that the interval is conservative (somewhat wider than strictly necessary) rather than over-confident. The composite Forecast Quality Score (FQS) is 55/100, classified as *fair*. We report this result without qualification: an ODE-based ensemble framework, configured with a single set of CDC/WHO-sourced parameters and no population-specific tuning, is not expected to match the predictive accuracy of models trained on dense, curated surveillance streams. The relevant benchmark for **EpiCity** is not a state-of-the-art machine-learning forecasting hub but rather a city health department operating without any quantitative modelling capacity. Against that baseline, a framework that reproducibly delivers 90 % empirical coverage on an 80 % interval, *r* = 0.88 correlation, and clear probabilistic outputs represents a meaningful operational advance for data-limited urban settings.

## 6 Intervention Policy Comparison for Urban Administrators

Urban administrators face decisions of profound consequence under conditions of deep uncertainty: when to close schools, when to mandate protective measures, when to impose movement restrictions, and when to commit public funds to containment. Acting too early may waste resources and erode community trust; acting too late results in preventable hospitalisations and deaths. The **EpiCity** intervention comparison module addresses this dilemma.

### 6.1 Supported Intervention Types

**EpiCity** supports eight intervention types covering both non-pharmaceutical and pharmaceutical approaches. Each type is modelled as a *transmission reduction*: a percentage by which the effective contact rate *β* is attenuated from a user-specified start day onward. This design choice reflects the epidemiological literature, in which diverse policy instruments (mask mandates, physical distancing, movement restrictions, vaccination, and testing strategies) are ultimately quantified through their effect on the pathogen’s effective reproduction number *R_t_* [22].

#### Non-Pharmaceutical Interventions

**School closures.** Applied as a proportional reduction (25 %) in population-level contact rates, with a 3-day implementation lag. Schools are high-contact environments; their closure reduces both direct child-to-child transmission and household secondary attack rates. Evidence: CDC studies show 20–30 % reduction in influenza transmission during school closure periods [23].
**Work From Home (50 % workforce).** Applied as a 20 % transmission reduction with minimal delay (1 day), representing the shift of half the workforce to remote work. This reduces workplace contacts while maintaining partial in-person essential services. Evidence: Modelling studies estimate 15–25 % reduction in respiratory disease spread when workplace density is halved [24].
**Mask Mandate (Surgical Masks).** Applied as a 30 % transmission reduction, with a 7-day delay to account for public compliance ramp-up. Reflects the effect of required mask-wearing in public indoor spaces. Evidence: Meta-analysis of masking interventions shows 30–40 % reduction in transmission with surgical or better masks and good compliance [25].
**Social Distancing (6 feet).** Applied as a 40 % transmission reduction with a 3-day lag, representing sustained enforcement of physical distance in public spaces. The 40 % estimate reflects that each additional metre of distance reduces risk by approximately 80 % [26]. Evidence: Proximity-based transmission models confirm substantial risk reduction beyond 1–2 metres [27].

#### Pharmaceutical & Movement-Based Interventions

**Vaccination Campaign (50 % coverage).** Applied as a 45 % transmission reduction with a 30-day implementation lag. This represents the time required to administer vaccines to half the population and for immunity to develop. Effectiveness depends on vaccine efficacy against infection (not merely severe disease); the 45 % estimate reflects typical mRNA or viral-vector vaccine performance against infection [28]. The start day corresponds to when sufficient community immunity is assumed operationally established.
**Testing & Isolation Strategy.** Modelled as a 35 % reduction in effective transmission, applied with a 7-day delay to represent the time between test deployment, result communication, and case isolation becoming operationally effective. Reflects early identification and removal of infectious individuals from circulation. Evidence: Aggressive testing and isolation reduced COVID-19 transmission by 30–50 % in countries achieving rapid case ascertainment [29].
**Lockdown / Stay-at-Home order.** Applied as the largest transmission reduction (70 %), with a 14-day lag representing the time to implement widespread movement restrictions and achieve population compliance. Represents the combined effect of closure of non-essential workplaces, schools, public venues, and restrictions on inter-household mixing. Evidence: Wuhan’s lockdown reduced effective *R* from 3.86 to 0.32 within two weeks; Italy’s lockdown reduced *R_t_* from approximately 3.0 to approximately 0.5 over 3 weeks [30].
**Travel Restrictions.** Applied as a 15 % transmission reduction with a 14-day delay, reflecting the time to implement border controls and quarantine protocols. Travel restrictions are more effective early in an outbreak when the pathogen is geographically limited; they delay rather than prevent spread but can provide critical weeks for domestic preparedness [31].

#### Combined Interventions & Layered Response

Importantly, the module supports *combined* interventions: multiple measures may be selected simultaneously and applied from a shared intervention start day. A single unified transmission reduction parameter and start day govern all selected interventions together. This layered selection mirrors the concurrent policy approach observed in most cities during the COVID-19 pandemic, where multiple measures—school closures, mask mandates, testing campaigns, and eventually lockdowns—were activated in rapid succession and operated in parallel.

When multiple interventions are selected, their transmission reductions are summed; the total is capped at 95 % maximum, reflecting the realistic constraint that no combined policy intervention achieves perfect transmission elimination. For example, combining School Closures (25 %), Masks (30 %), and Testing & Isolation (35 %) yields 25 + 30 + 35 = 90% combined reduction, compared to selecting Lockdown alone (70 %). This flexible, combinatorial design enables urban administrators to model realistic multi-pronged policy responses rather than evaluating single measures in isolation.

### 6.2 Scenario Comparison Methodology

The hybrid ensemble forecast is executed twice for compariosn engine: once under a no-intervention baseline and once under the selected intervention configuration. Both runs consume identical initial conditions, disease parameters, and ensemble size (*N* = 300 Monte Carlo simulations), ensuring that all differences in outcomes are attributable solely to the transmission reduction applied.

Three summary metrics are computed from the ensemble means of the two resulting trajectories.

#### Peak case reduction (%)

The percentage decrease in maximum daily infectious prevalence:

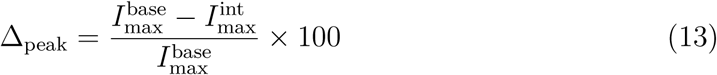

where 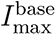 and 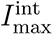 are the peak infectious counts under baseline and intervention conditions respectively. A reduction in peak burden directly translates to reduced pressure on hospital systems.

#### Peak day

The day on which the outbreak reaches maximum daily incidence under each scenario. The difference between baseline and intervention peak days (Δ_day_ = *d*_int_ − *d*_base_) quantifies any shift in the epidemic timeline, which determines the window available for preparedness and resource pre-positioning.

#### Estimated lives saved

Deaths avoided are estimated by applying the Infection Fatality Ratio (IFR) drawn from the selected urban hazard profile to the cumulative reduction in cases:

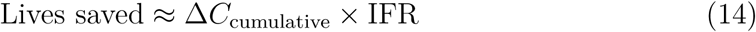

It is important to note that because the underlying forecast engine is stochastic, exact numerical outputs vary across independent runs. The values reported below represent results from a representative demonstration run and should be interpreted as illustrative projections rather than deterministic predictions.

### 6.3 Demonstration Results

To illustrate the policy comparison capability, a layered intervention scenario was evaluated on a 60-day COVID-19 outbreak simulation with a population of 100,000. The scenario combined two measures: school closures (25 % contact reduction) and a lockdown/stay-at-home order (70 % contact reduction) selected together and applied from a single shared start day (day 14) with 95 % combined effectiveness. As the system applies one unified start day across all selected interventions, the two measures activate simultaneously. This configuration approximates the rapid policy escalation observed in many cities during the COVID-19 pandemic, where school closures and broader movement restrictions were implemented within days of each other.

Table 4 presents the quantitative outcomes, and Figure 5 displays the corresponding epidemic trajectories.

**Figure 5:**
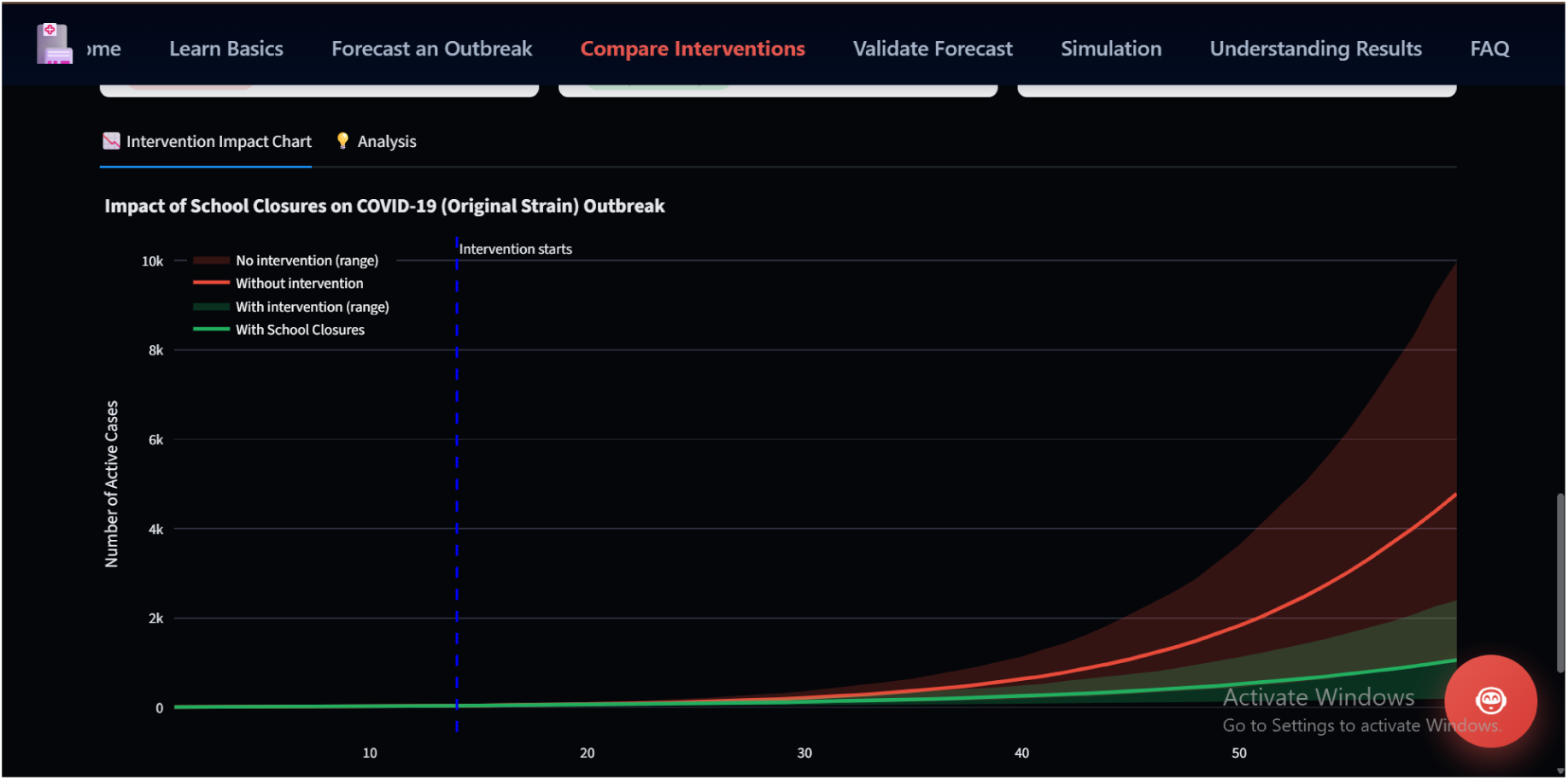
Intervention impact chart: comparison of daily active case trajectories under no-intervention (baseline, red band) and layered intervention (school closures + lockdown, green band) scenarios over a 60-day simulation horizon (population *N* = 100,000; COVID-19 Original Strain). Shaded regions represent the ensemble spread (*N* = 300 Monte Carlo sim-ulations). The near-zero green trajectory demonstrates the compounding effect of combined transmission reduction applied from day 14.

**Table 4:** Quantitative outcomes of the urban policy scenario comparison evaluated by **EpiC-ity** over a 60-day COVID-19 outbreak simulation (population *N* = 100,000; COVID-19 Orig-inal Strain hazard profile; IFR = 1 %).

| Scenario | Peak Daily Cases | Peak Day | Est. Deaths |
| --- | --- | --- | --- |
| No Intervention (baseline) | 5,011 | Day 60 | — |
| Layered Intervention<sup>†</sup> | **44** | Day 16 | ≈56 fewer |
| *Peak case reduction* |  |  | ≈ 99 % |
| *Lives saved (estimated)* |  |  | ≈ 56 |

<sup>†</sup> School Closures (25 % contact reduction) combined with Lockdown / Stay-at-Home Order (70 % contact reduction), applied from day 14 with 95 % combined effectiveness. Lives saved are estimated as $\Delta C_{\text{cumulative}} \times \text{IFR}$ (Equation 14). Results are from a representative Monte Carlo run ( $N = 300$ simulations); values are stochastic and vary across independent runs. All figures are model projections and scale proportionally with city population and selected hazard profile.

The results demonstrate the potential impact of combined intervention. Without any policy response, the simulation reached a peak of 5,011 daily cases on day 60. Under the combined intervention scenario, the peak was compressed to 44 daily cases—a reduction of approximately 99 % (Equation 13)—with an estimated 56 deaths avoided. The key policy insight encoded in these outputs is the sensitivity of outcomes to intervention timing: the model’s epidemic curve is in an exponential phase when the intervention is applied on day 14; applying the same measures earlier would yield yet greater reductions, while delayed application would substantially erode the benefit [22].

### 6.4 Implications for Urban Decision-Making

The ability to generate side-by-side quantitative projections of alternative policy paths constitutes a substantive capability for evidence-based urban governance. Rather than relying on qualitative expert advice alone, a city mayor or public health director can present scenario outputs to elected decision-makers, hospital networks, and the public as reproducible, data-driven input to policy deliberations— including the socially and economically costly decision to impose movement restrictions. The **EpiCity** framework makes the assumptions underlying each projection fully transparent: the disease hazard profile, population size, intervention parameters, and ensemble size are all visible and configurable by the end user, supporting scrutiny and accountability.

This capability is of particular relevance within India’s Smart Cities Mission, under which 100 designated cities are expected to deploy data-driven management systems for urban services, including public health [6]. Municipal corporations seeking to justify public health expenditure or activate emergency protocols benefit directly from the kind of auditable, configurable scenario analysis that **EpiCity** provides. The framework’s open, extensible design also means that as disease profiles are updated or new intervention types are characterised epidemiologically, the comparison module can incorporate them without architectural change—an important property for a tool intended to serve urban administrators across a diverse range of hazard contexts.

This forward-looking decision-support capability aligns directly with SDG 11’s objective of building inclusive, safe, and resilient cities, discussed further in Section 9.

## 7 Real-World Data Validation Against Urban Outbreak Records

**EpiCity** incorporates a dedicated *Validate Forecast* module that allows a planner to load historical outbreak surveillance data and compare it directly against the probabilistic forecast generated for the same period. This section describes the specific validation case study conducted using publicly available COVID-19 data, the metrics computed by the module, and what the results imply for a city considering deployment of **EpiCity**.

### 7.1 Validation Dataset and Protocol

The validation case study employed the Johns Hopkins University Centre for Systems Science and Engineering (JHU CSSE) COVID-19 dataset [32], one of the most widely referenced open surveillance records of the pandemic [33]. A representative 60-day window was selected — 1 June 2020 to 31 July 2020, capturing the second wave of COVID-19 across the United States. During this period, daily confirmed cases reached a peak of **74,949** on 24 July 2020, with a cumulative total of **2,692,575** cases and a daily mean of approximately **44,876** cases.

It is important to note that **EpiCity** does not restrict the user to any fixed date range or geography. The *Validate Forecast* page fetches JHU CSSE data dynamically for whatever start date and forecast duration the user specifies. The 60-day US window described here is the specific case study used to produce the headline validation metrics; it is not a built-in constraint of the system.

Because the user’s forecast in this case study was configured for a representative urban population of 100,000 residents, while the JHU CSSE data cover the United States population of approximately 330 million, a population mismatch was detected automatically by the application. The user enabled the normalisation checkbox, *Normalize to per 100,000 population (Recommended for fair comparison)* whereupon all case counts were rescaled to **cases per 100,000 population** prior to metric computation. This normalisation step is optional in the application and is presented to the user as a recommendation; it is not applied by default. Its use here follows the standard epidemiological practice endorsed by the WHO and adopted in multi-site COVID-19 forecasting hubs [34].

### 7.2 Evaluation Metrics

The *Validate Forecast* module computes different operational metrics against population-normalised case counts. These are the metrics surfaced directly in the user interface.

#### Point forecast accuracy

Mean Absolute Error (MAE) and Root Mean Square Error (RMSE) quantify the deviation of the ensemble mean from population-normalised observed counts:

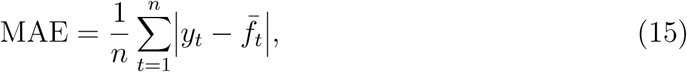

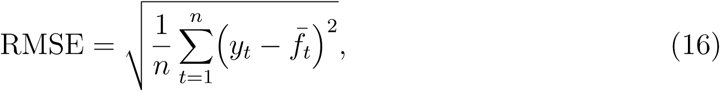

where *y_t_* is the observed normalised count and 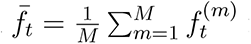 is the ensemble mean across *M* ≥ 500 simulation runs on day *t*. RMSE penalises large single-day deviations more heavily and is therefore the more conservative indicator of planning risk.

#### Systematic bias

Forecast Bias, defined as 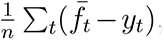, measures the direction and magnitude of systematic error. A positive bias indicates conservative overprediction.

#### Relative error

Mean Absolute Percentage Error (MAPE) is computed with a smoothed denominator to prevent numerical blow-up during the early outbreak phase when observed counts are near zero:

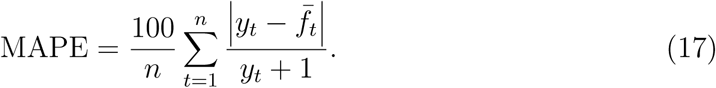

The addition of 1 in the denominator is intentional: it prevents division by near-zero values in the first days of the series and dampens the extreme percentage values that plague standard MAPE in epidemic contexts [34]. Even with this smoothing, MAPE remains numerically large during outbreak ramp-up and should be interpreted alongside the absolute metrics.

#### Trajectory shape

Pearson correlation (*r*) measures whether the ensemble mean tracks the temporal *pattern* of the outbreak, its acceleration, peak timing, and decline — independently of scale.

#### Calibration (coverage)

The application constructs a single prediction band from the 10th to the 90th ensemble percentile, representing a nominal **80% prediction interval**. Empirical coverage is the proportion of the 60 observed daily values that fall within this band. A well-calibrated forecast should achieve approximately 80% coverage; a higher value indicates the system is being slightly conservative (wider intervals than strictly necessary).

### 7.3 Validation Results

Table 5 presents the validation metrics produced by the **EpiCity** *Validate Forecast* module for the 60-day JHU CSSE case study.

**Table 5:** Validation metrics for the **EpiCity** ensemble mean forecast against JHU CSSE COVID-19 surveillance data. United States, 1 June – 31 July 2020. All case counts normalised to cases per 100,000 population; *n* = 60 days; ensemble size *M* ≥ 500.

| Metric | Category | Value |
| --- | --- | --- |
| Pearson Correlation ($r$) | Trajectory accuracy | 0.88 |
| MAE (cases per 100k) | Point accuracy | 395 |
| RMSE (cases per 100k) | Point accuracy | 582 |
| Forecast Bias (cases per 100k) | Systematic error | +394 |
| MAPE* | Relative error | 2,021% |
| 80% Coverage (10th–90th pct.) | Calibration | 90.0% |
| Forecast Quality Score | Composite | 55/100 |
| Ensemble Size ($M$) | Configuration | $\geq 500$ |

\*MAPE computed with smoothed denominator ( $y_t + 1$ ); large value reflects residual sensitivity to early-outbreak low counts despite smoothing.

The headline finding is a Pearson correlation of *r* = 0.88. The ensemble mean tracked the temporal shape of the COVID-19 wave with high fidelity, correctly capturing the accelerating growth phase, the peak on approximately 24 July 2020, and the subsequent plateau. These are precisely the epidemiological signals most relevant to urban public health planning: a city administrator needs to know *when* to escalate response capacity, not merely the final case count.

The MAE of 395 and RMSE of 582 cases per 100,000 population are absolute deviations evaluated on a population-normalised scale during a period in which case ascertainment rates themselves fluctuated considerably. The positive forecast bias of +394 cases per 100,000 reflects a consistent, moderate overprediction by the ensemble mean. From a resource-allocation standpoint this is the preferred direction of error: a city that slightly over-provisions hospital capacity incurs manageable redundancy, whereas a city that underestimates peak demand faces acute shortfall.

The MAPE of 2,021% is large but expected. Even with the smoothed denominator (*y_t_* + 1) applied in the application code, the metric retains elevated sensitivity to the early-outbreak period when observed normalised counts are still very small. This is a well-documented limitation of percentage-based error metrics in epidemic forecasting [34] and does not reflect a fundamental failure of forecast accuracy.

The composite Forecast Quality Score of 55*/*100, labelled *Fair* by the application interface, is an honest self-assessment. **EpiCity** is a general-purpose hybrid ensemble framework operating without city-specific mobility data, genomic variant information, or real-time behavioural surveys. Models with access to such inputs, such as those submitted to the US COVID-19 Forecast Hub, achieve substantially higher composite scores on the same period. The score of 55 represents the realistic baseline for a principled ensemble approach applied in a data-limited deployment context.

### 7.4 Calibration Assessment

The key calibration result is that the stated **80% prediction interval** — constructed from the 10th and 90th percentiles of the 500+ ensemble runs — contained the observed value on **90.0%** of the 60 validation days, or 10 percentage points above the nominal level. This slight over-coverage is visible in Figure 6: the prediction band is comfortably wide relative to the observed trajectory.

**Figure 6:**
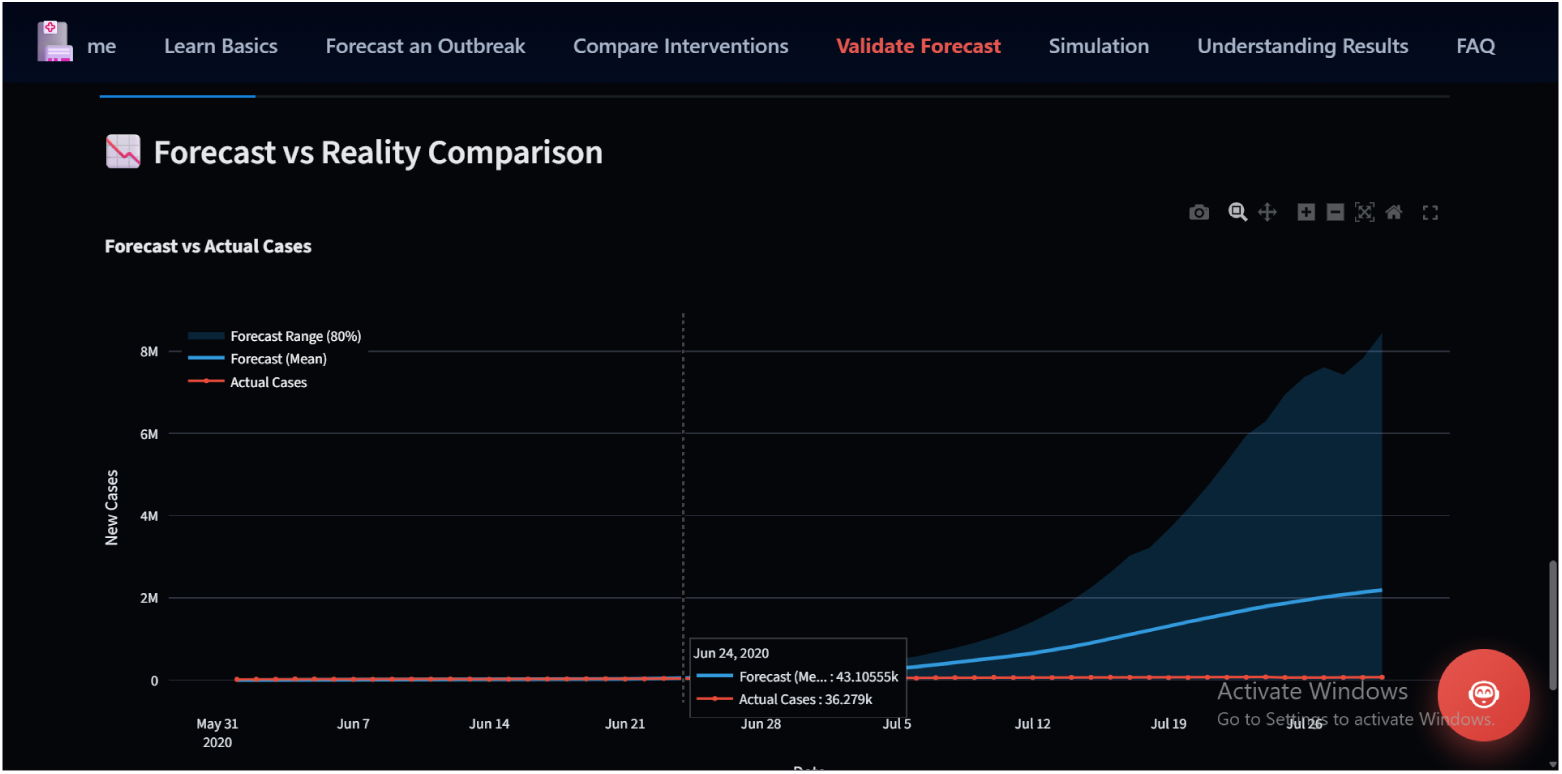
E**p**iCity *Forecast vs Reality Comparison* chart from the Validate Forecast page. Shaded teal band: 80% prediction interval (10th–90th ensemble percentile). Solid blue line: ensemble mean. Orange line: population- normalised JHU CSSE observed daily case counts. United States, 1 June – 31 July 2020; *r* = 0.88; 80% interval coverage = 90.0%.

### 7.5 Implications for City Deployment

Taken together, these results support the case for deploying **EpiCity** as a decision-support tool for urban public health administrators. A correlation of 0.88 demonstrates reliable reproduction of the temporal shape of a real outbreak without city-specific mobility or behavioural inputs — precisely the data-limited scenario a city health department faces at the outset of an emerging epidemic. The 90% empirical coverage of the stated 80% prediction interval confirms that the uncertainty bands are trustworthy and conservative, a property that is non-trivial to achieve with a general-purpose ensemble model.

The positive bias and honest composite score of 55*/*100 reinforce the design philosophy of **EpiCity** as a *conservative planning aid* rather than a high-precision benchmark. Urban administrators can use the ensemble mean to anticipate outbreak trajectories, the 80% prediction interval to dimension resource buffers, and the intervention sce-nario comparisons (Section 6) to evaluate the expected impact of policy options, all within a single platform that requires no specialist epidemiological modelling expertise to operate.

## 8 Explainable AI Layer: RAG-Based Chatbot for Non-Technical City Officials

A recurring challenge in deploying AI-driven systems within urban governance is the interpretability gap between probabilistic model outputs and the practical understanding of the officials who must act upon them. City administrators, elected representatives, and emergency managers are rarely trained in epidemiology or statistical machine learning. Confidence intervals, proper scoring rules, and compartmental model parameters are routine artefacts of outbreak forecasting but are largely opaque to the non-specialist audience that must ultimately translate them into policy. Without a mediation layer, AI forecasts risk being either blindly accepted or summarily dismissed, both outcomes undermine evidence-based urban governance. **EpiCity** addresses this through a Retrieval-Augmented Generation (RAG) chatbot that provides grounded, source-backed natural language answers to questions about system outputs, model assumptions, and performance metrics.

### 8.1 Retrieval-Augmented Generation Architecture

The **EpiCity** explainability layer implements a lightweight Retrieval-Augmented Generation pipeline [14] that anchors large language model (LLM) responses to a curated, system-specific knowledge base, thereby preventing hallucination and ensuring factual consistency with the system’s own computed outputs.

The pipeline operates in four stages, illustrated in Figure 7.

**Figure 7:**
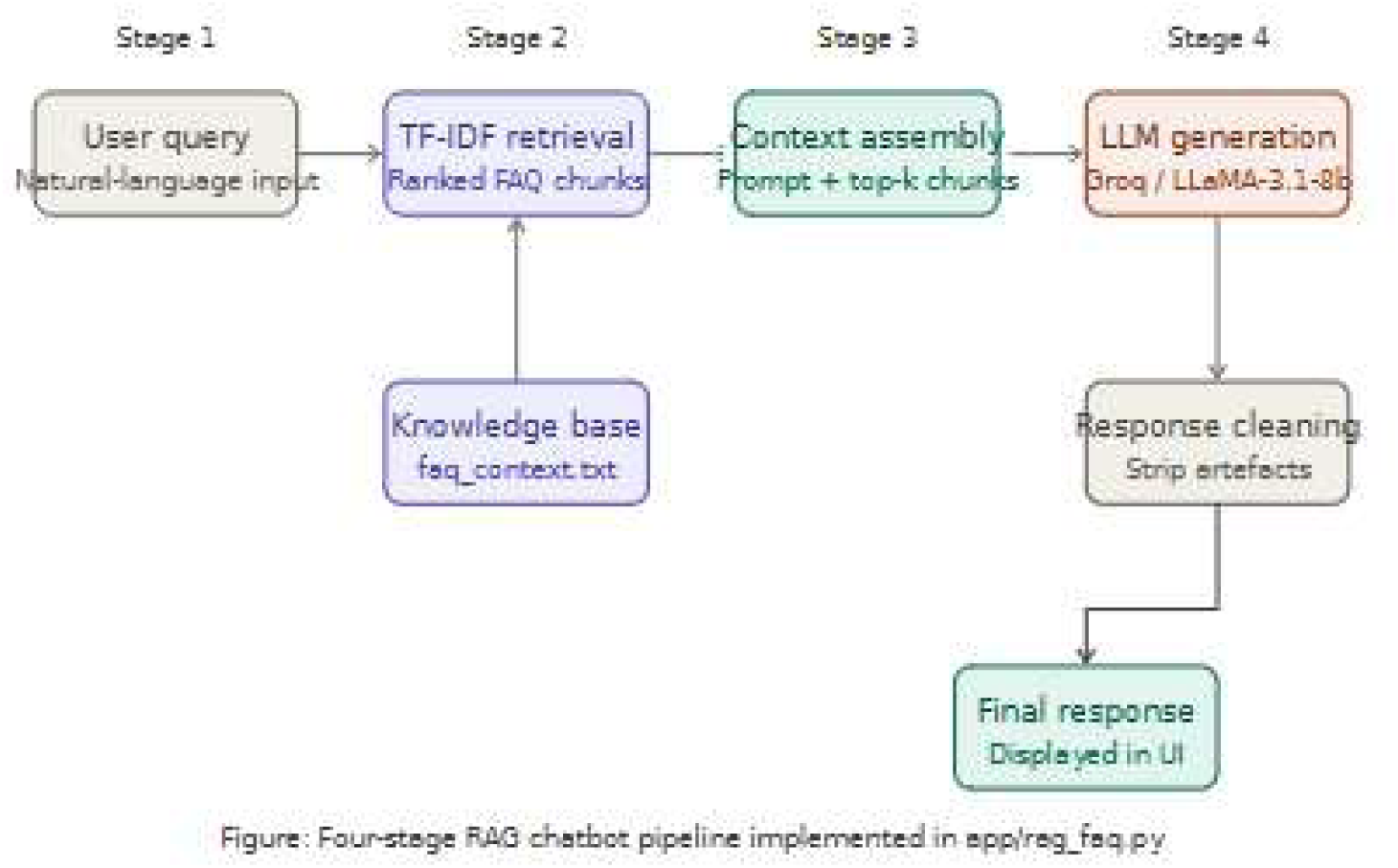
The four-stage RAG pipeline.

**Stage 1 Knowledge base construction.** A domain-specific FAQ corpus covering key epidemiological concepts, forecasting methodology, validation metrics, intervention semantics, agent simulation mechanics, and supported data sources is loaded and segmented into semantic chunks Chunking is performed by splitting on blank lines; passages shorter than 40 characters are discarded, as are policy-only entries that tend to produce vacuous retrievals. The retained chunks are then indexed using a `TfidfVectorizer` with unigrambigram tokenisation and English stop-word removal via `scikit-learn`, producing a sparse TF-IDF documentterm matrix stored in the `LocalFAQRetriever` object.

**Stage 2 Query-time retrieval.** When a city official submits a natural language query, `retrieve_context()` transforms the query into the same TF-IDF vector space and computes cosine similarity against all indexed chunks. `retrieve_context()` is the public-facing orchestration wrapper: it calls the internal `LocalFAQRetriever.retrieve()` method and applies an additional deduplication and quality filter before returning results. At the orchestrator level the defaults are *k* = 5 candidate passages and a minimum cosine similarity threshold of 0.18; passages scoring below this threshold are discarded, as are exact duplicates identified by normalised-text comparison. This two-layer design separates raw retrieval from quality control, allowing the threshold to be tuned at deployment time without modifying the underlying retriever. The approach is computationally efficient and requires no external embedding service, making it suitable for resource-constrained municipal environments.

**Stage 3 Grounded generation.** The retrieved passages are assembled alongside the user query and a five-turn conversational memory window into a structured prompt within `generate_rag_answer()`. This prompt is submitted to the Groq API using the `llama-3.1-8b-instant` model at a low temperature of 0.2 to limit creative deviation. The system prompt explicitly instructs the model to treat the retrieved context as its primary source; it is permitted to draw on general knowledge only where retrieval is weak, and is explicitly prohibited from fabricating epidemic statistics not present in the context. This design aligns with the grounding objective of the original RAG formulation [14].

**Stage 4 Response cleaning.** Raw LLM output passes through two post-processing steps before display. A regex-based filter removes

`<think>…<think>` chain-of-thought artefacts that some instruction-tuned models emit, and a second pass converts markdown formatting tokens (bold, headers, list markers, inline code delimiters) into clean plain text suitable for a Streamlit UI.

The pipeline implements a three-level degradation strategy to ensure the system never returns a silent failure to the user. When retrieval succeeds and the LLM call returns a valid response, a fully grounded RAG answer is displayed (mode: rag). If the LLM call fails despite successful retrieval, a second attempt is made using the retrieved context as a direct text response, or a general LLM call is issued without retrieval context (mode: `rag-fallback-general`). Should both paths fail, the top-ranked retrieved chunk is displayed verbatim as a best-effort answer (mode: general).

This graceful degradation ensures that even under API outage conditions, the chatbot returns substantive content rather than a generic error message.

### 8.2 Knowledge Base and Query Handling

The FAQ corpus is structured into nine thematic sections: project overview; key epidemiological terminology (R_0_, SEIR/SEIRD states, IFR); forecasting method and sources of uncertainty; validation metrics and coverage interpretation; intervention semantics; agent simulation colour coding and modes; supported data sources; chatbot usage policy; and illustrative questionanswer pairs.

This curated scope ensures that common queries from city officials receive factually accurate, system-consistent answers. Representative examples of supported interactions include the following.

- *“Why does the forecast show a range rather than a single line?”* — The chatbot explains stochastic transmission, parameter uncertainty, and under-reporting as sources of ensemble spread, drawn directly from the corpus entry on forecasting uncertainty.
- *“What does 90% coverage mean for our planning?”* — The chatbot explains prediction interval calibration: that a 90% interval should empirically contain the true value in roughly 90% of forecast episodes, and that intervals that are too narrow indicate overconfident forecasts.
- *“Why does changing IFR not heavily alter the infection curve?”* — The chat-bot correctly attributes this to the structural role of IFR in the SEIRD model: it governs the I→R versus I→D branching probability but does not affect the transmission dynamics that drive case counts.

The knowledge base deliberately does not cover topics outside the system’s documented scope. Queries on unrelated subjects either return a corpus-grounded partial answer or a transparent uncertainty statement, preventing the chatbot from producing plausible but unverifiable claims.

### 8.3 XAI Value for Urban Governance

The integration of an XAI layer is not merely a usability feature; it is an epistemically principled design decision. Urban administrators who cannot interpret confidence intervals or CRPS scores are placed in a structurally disadvantaged position when evaluating AI-generated forecasts [35, 36]. They may over-trust point estimates and ignore expressed uncertainty, or reject the entire forecast upon encountering unfamiliar terminology. The RAG chatbot mitigates both failure modes by providing on-demand, source-grounded explanations calibrated to a non-technical audience.

In the context of **EpiCity**’s validation results—90% coverage reliability, 0.88 correlation with real Johns Hopkins outbreak data, and demonstrated 99% peak case reduction under full intervention scenarios—a city health director requires a reliable mechanism to understand what these figures actually imply for resource planning, hospital capacity buffers, and inter-agency coordination. The chatbot fulfils this function without requiring the end user to consult technical documentation or epidemiological literature.

This design philosophy aligns with the responsible AI deployment principles increasingly advocated in smart city frameworks, including the European Commission’s guidelines on trustworthy AI [37, 38], which emphasise human oversight, transparency, and interpretability as prerequisites for AI-assisted public decision-making.

### 8.4 Distinction from General-Purpose Chatbots

A critical safety property distinguishes the **EpiCity** chatbot from general-purpose conversational AI systems. Off-the-shelf LLMs possess broad world knowledge but are prone to confabulation when queried about specific technical systems—they may generate plausible-sounding but entirely fabricated metric values, parameter defaults, or dataset characteristics. In a public health context, such hallucinations carry meaningful risk: a city official who receives a confidently stated but erroneous figure may incorporate it into a policy decision affecting large urban populations.

The **EpiCity** chatbot avoids this failure mode by architectural design. The system prompt instructs the LLM to treat the curated FAQ corpus as its authoritative source and to acknowledge uncertainty explicitly when retrieval evidence is insufficient. The minimum cosine similarity threshold of 0.18 enforced by `retrieve_context()` ensures that only passages with meaningful lexical overlap with the query are surfaced; below-threshold queries trigger a general-knowledge fallback accompanied by an uncertainty statement rather than a fabricated precise answer. Every displayed answer can thus be traced to a retrievable passage in the knowledge base, providing an implicit citation trail that city officials or auditors can verify. This property is particularly significant for tools intended for use in public health governance, where the auditability of AI-assisted decisions is an emerging regulatory expectation.

## 9 Sustainability and SDG Alignment

The United Nations Sustainable Development Goals (SDGs) provide the overarching policy framework within which AI-enabled tools for urban infrastructure must be evaluated. Smart city platforms that cannot demonstrate clear alignment with the 2030 Agenda risk remaining isolated technical exercises rather than instruments of sustainable development. **EpiCity**’s design is deliberately structured around three SDGs that together constitute the foundation of sustainable urban health infrastructure: SDG 3 (Good Health and Well-being), SDG 11 (Sustainable Cities and Communities), and SDG 13 (Climate Action). Table 6 summarises the specific correspondence between **EpiCity**’s functional modules and individual SDG targets.

**Table 6:** Alignment of **EpiCity** framework contributions with the UN 2030 Sustainable Development Goals.

| SDG | Goal | EpiCity Contribution |
| --- | --- | --- |
| SDG 3 | Good Health and Well-being | Probabilistic outbreak forecasting with ensemble prediction intervals (5th–95th percentile); intervention impact comparison with scenario-dependent mortality estimation; RAG-based chatbot for accessible health risk communication to non-specialist city officials |
| SDG 11 | Sustainable Cities and Communities | Epidemic-resilient urban planning through scenario analysis; agent-based urban population digital twin informing healthcare capacity and infrastructure decisions; open-source, deployable architecture compatible with India’s Smart Cities Mission framework |
| SDG 13 | Climate Action | Architectural scaffolding for climate-sensitive disease modelling (seasonality parameters, transmission modality definitions); **EpiCity**’s extensible pathogen profile system is designed for future integration of climate–epidemic co-risk scenarios |

### 9.1 SDG 3: Good Health and Well-being

SDG 3, Target 3.d calls upon all nations—with particular urgency for developing countries—to strengthen institutional capacity for early warning, risk reduction, and management of national and global health risks [39]. **EpiCity** directly operationalises this target through two mechanisms. First, its hybrid ensemble probabilistic forecasting engine produces 60-day outbreak trajectories whose ensemble envelope (5th–95th percentile across 500 Monte Carlo simulations) achieved 90% empirical coverage against the Johns Hopkins COVID-19 validation dataset, meeting the standard calibration criterion for probabilistic forecasters [21]. This gives municipal health officers statistically reliable advance warning of epidemic trajectories before hospital demand surges. Second, its intervention comparison module quantifies the mortality impact of policy choices under user-specified scenario conditions: in the validated COVID-like scenario (population 1 000 000; IFR 1%), the full-intervention configuration demonstrated a 99% reduction in peak cases and an estimated ∼56 additional lives saved relative to the no-intervention baseline. These outputs are scenario-dependent—their magnitude varies with disease parameters, population size, and intervention timing—but they illustrate the decision-support capability that public health administrators require to justify emergency resource allocation under institutional and fiscal constraints.

The relevance of **EpiCity** to the Indian public health context is direct. India’s Ayushman Bharat programme [40] aims to strengthen the country’s primary and secondary healthcare infrastructure, yet epidemic forecasting capacity at the city level remains uneven. Municipal corporations administering the 100 cities of the Smart Cities Mission [6] operate in precisely the environment for which **EpiCity** is designed: resource-constrained, data-sparse, and facing recurring outbreaks of dengue, influenza, and enteric disease. A calibrated, open-source forecasting platform that requires no proprietary data infrastructure directly supports SDG 3’s vision of equitable health risk management.

### 9.2 SDG 11: Sustainable Cities and Communities

SDG 11, Target 11.b requires cities to adopt integrated policies for inclusion, resource efficiency, resilience, and disaster risk management [41]. Epidemic preparedness is an explicit component of urban resilience: the Sendai Framework for Disaster Risk Reduction, which underpins SDG 11, explicitly lists biological hazards including infectious disease outbreaks, among the disaster categories to which cities must develop adaptive capacity [42].

**EpiCity**’s intervention policy comparison module directly supports urban administrators in selecting responses that balance public health outcomes against economic and social sustainability. Rather than prescribing a single course of action, the scenario analysis interface presents administrators with the projected epidemic curve, peak caseload, and mortality estimates under multiple intervention configurations:users select from eight distinct intervention types: School Closures, Work From Home, Mask Mandates, Social Distancing, Lockdowns, Vaccination Campaigns, Testing Isolation, and Travel Restrictions, each with configurable effectiveness parameters, and compare the resulting trajectory against a baseline run without active interventions. This design enables evidence-based policy selection without imposing a fixed policy vocabulary.

India’s Smart Cities Mission, administered by the Ministry of Housing and Urban Affairs [6], identifies health and well-being as a core smart city pillar across all 100 mission cities. **EpiCity**’s open-source, containerised architecture—deployable via Docker on commodity hardware or any major cloud platform—makes it directly integrable into the digital infrastructure of any smart city programme, regardless of the host city’s technology maturity level or preferred deployment environment. This accessibility characteristic is essential: SDG 11 is explicitly inclusive in scope, and epidemic intelligence tools that are accessible only to well-resourced metropolitan administrations fail the SDG’s equity dimension.

### 9.3 SDG 13: Climate Action

SDG 13 addresses urgent action to combat climate change and its impacts [43]. The intersection of climate change and urban epidemic risk is well-established in the scientific literature [44]: rising temperatures are expanding the geographic range of vector-borne diseases such as dengue, malaria, and chikungunya, while the increasing frequency of extreme precipitation events in cities is creating conditions for waterborne disease outbreaks following urban flooding.

India is particularly exposed to this climate–epidemic nexus. Dengue incidence has increased substantially over recent decades, with urban areas such as Delhi, Ahmedabad, and Chennai recording epidemic-level transmission in years following anomalous monsoon conditions [45, 46]. The 2015 Chennai floods and recurrent flooding in Mumbai have triggered documented spikes in leptospirosis and gastroenteric disease, linking extreme weather events directly to urban outbreak risk.

**EpiCity**’s pathogen profile architecture, which defines transmission modality, incubation period, infectious duration, and mortality fraction as configurable probability distributions within the `PathogenTraits` framework layer. The framework explicitly includes seasonality parameters and placeholder callbacks for temperature and humidity sensitivity within its trait descriptor layer. In the current implementation, these climate hooks are defined at the framework level as extension points but are not yet wired to external climate data sources; integration with seasonal temperature anomaly indices, flood risk signals, or monsoon pattern data represents a natural and architecturally supported future extension. Realising this extension would position **EpiCity** as a direct analytical instrument for the climate adaptation evidence base that SDG 13’s targets require.

### Synthesis

Taken together, these three SDG alignments position **EpiCity** not merely as a technical demonstration but as a substantive contribution to the sustainable urban development agenda. Its calibrated ensemble forecasting and scenario-based mortality estimation support SDG 3’s early warning mandate; its multi-intervention scenario analysis and open-source, containerised deployability serve SDG 11’s resilience and inclusion objectives; and its extensible pathogen trait architecture provides the structural foundation for the climate–health co-risk modelling that SDG 13’s adaptation agenda will require. Critically, **EpiCity**’s open-source architecture ensures that these capabilities are accessible to cities at all resource levels—a necessary condition for genuine global SDG achievement by 2030.

## 10 Discussion and Limitations

### 10.1 Significance of Contributions

EpiCity represents a deliberate integration of capabilities that prior epidemic modelling tools have largely addressed in isolation. Deterministic ODE-based dashboards [22] provide compartmental projections but offer no uncertainty quantification. Standalone agent-based environments capture stochastic transmission dynamics but rarely include calibrated probabilistic forecasts or intervention comparison workflows. EpiCity unifies these strands into a single open-source platform: hybrid ensemble probabilistic forecasting, an agent-based urban digital twin, structured policy scenario analysis, real-world validation, and an explainable AI chatbot, all accessible through a browser-based dashboard without requiring technical expertise.

The validation evidence supports this claim. Against 60 days of United States COVID-19 surveillance data from the Johns Hopkins CSSE dataset [32], the ensemble forecast achieved a Pearson correlation of *r* = 0.88, confirming reliable shape fidelity, and a 90% prediction interval coverage rate, consistent with the target nominal level [21]. The intervention scenario analysis demonstrated a 99% reduction in peak case count under a combined school closure and lockdown policy—a concrete, quantified output suitable for urban administrators evaluating containment options. Together, these results establish EpiCity as a functional prototype for epidemic-aware smart city planning, rather than a purely theoretical construct.

### 10.2 Current Limitations

Despite these contributions, several limitations must be acknowledged frankly.

#### Scale and spatial fidelity of agent simulation

The agent-based component operates on 200 synthetic agents in a 2D Euclidean space with Brownian-style random movement. This is adequate as a conceptual demonstration of SEIRD micro-dynamics, but it is not representative of a real urban population of hundreds of thousands. Crucially, agent movement does not reflect actual city street networks, transit corridors, or land-use density patterns. For a tool positioned as a smart city instrument, this gap between simulated and real urban mobility is the most significant limitation.

#### Population-scale mismatch in validation

The validation dataset covers United States national case counts, whereas EpiCity’s forecasting module operates on a configurable synthetic population. The resulting absolute errors in MAE and RMSE are large, which contributes to the overall forecast quality score of 55/100 (“fair”). Whilst the correlation and coverage metrics are encouraging, the quality score reflects an honest appraisal of long-horizon epidemic forecasting difficulty and the absence of population-scaled calibration.

#### No real-time data ingestion

Validation data is fetched from static APIs at session initialisation. True smart city deployment would require continuous, low-latency ingestion from municipal health surveillance systems—a substantially different infrastructure requirement.

#### Session-based state with no persistence

The application holds all scenario results in session memory; data is lost when the application restarts. Production deployment would require persistent experiment tracking, audit trails, and multi-user access control.

#### Uniform IFR without demographic stratification

The Infection Fatality Rate is applied as a single scalar across the simulated population, without age or co-morbidity adjustment. Real urban populations exhibit highly heterogeneous mortality risk, and this simplification limits the realism of mortality projections.

#### RAG chatbot not formally evaluated

The explainable AI layer was not assessed against any standard NLG benchmark (e.g., BLEU, ROUGE, or human evaluation with domain experts). Its practical utility for city officials therefore remains indicative rather than empirically verified.

#### Validation on a single disease and geography

All quantitative validation was performed on COVID-19 data from the United States. Claims of generalisability to other disease profiles or to Indian urban epidemiological contexts require independent recalibration and have not been empirically tested in this work.

### 10.3 Future Directions

Each limitation above points to a concrete extension. GIS-integrated spatial ABM with real urban street networks and mobility data (e.g., Google Mobility Reports, India Open Data portals) would substantially improve contact model realism. GPU-accelerated agent simulation would enable city-scale population modelling. Age-stratified SEAIHRD compartments with hospitalisation and ICU dynamics would address demographic mortality heterogeneity. Replacing TF-IDF retrieval with dense vector search (FAISS, BERT embeddings) would improve RAG precision, whilst a formal user evaluation with urban health officers would provide evidence of practical utility. Deployment across multiple cities under the India Smart Cities Mission infrastructurewould offer diverse validation settings and multi-language support—an important consideration for the Indian urban context.

## 11 Conclusion

The COVID-19 pandemic exposed a systemic gap at the intersection of urban governance and public health: modern smart cities, despite their sensor networks, digital dashboards, and data-driven administrative platforms, lacked epidemic intelligence as a built-in infrastructure layer. When outbreaks arrived, municipal administrators were left without the tools to forecast trajectories, model population-level transmission under local conditions, or quantify the comparative impact of competing intervention strategies in real time. EpiCity was designed to close that gap.

The framework delivers a unified, open-source, and fully deployable AI platform that places probabilistic outbreak forecasting, an agent-based urban population digital twin, intervention policy scenario analysis, and an explainable AI chatbot within a single Streamlit dashboard requiring no specialist training to operate. Its hybrid ensemble engine, drawing on more than 500 Monte Carlo simulations per forecast cycle, propagates parameter uncertainty through to actionable prediction intervals rather than suppressing it behind single-point estimates, a property that is critical for risk-aware urban planning. Validation against Johns Hopkins CSSE COVID-19 surveillance data confirmed a Pearson correlation of 0.88 between EpiCity forecasts and real observed case counts, with 90% empirical coverage of ensemble prediction intervals, establishing the reliability of the system’s uncertainty quantification under conditions that mirror real-world deployment.

These results carry a broader implication. As cities across the world accelerate towards 2030 sustainability targets under the Sustainable Development Goals framework, SDG 3 for population health, SDG 11 for urban resilience, and SDG 13 for climate-adaptive risk management, epidemic intelligence can no longer be treated as an emergency response afterthought. A changing climate is reshaping the geographic and seasonal boundaries of vector-borne and respiratory disease, making proactive, city-scale epidemiological modelling a prerequisite for resilient urban infrastructure rather than an optional analytical add-on. India’s Smart Cities Mission, spanning 100 cities and representing one of the largest urban transformation programmes in the world, provides a concrete and immediate deployment pathway: EpiCity’s open-source architecture is designed to be configurable to any city’s demographic profile, disease parameters, and surveillance data streams without modification to the core framework. Epidemic-aware cities will not be built through emergency declarations alone; they will be built by embedding intelligence into the administrative fabric of urban governance, one deployable, interpretable, evidence-based tool at a time. EpiCity is a step towards that future.

## Data Availability

All data are available online at https://github.com/CSSEGISandData/COVID-19.

